# What do we know about SARS-CoV-2 transmission? A systematic review and meta-analysis of the secondary attack rate, serial interval, and asymptomatic infection

**DOI:** 10.1101/2020.05.21.20108746

**Authors:** Wee Chian Koh, Lin Naing, Muhammad Ali Rosledzana, Mohammad Fathi Alikhan, Liling Chaw, Matthew Griffith, Roberta Pastore, Justin Wong

**Affiliations:** Centre for Strategic and Policy Studies, Brunei Darussalam; PAPRSB Institute of Health Sciences, Universiti Brunei Darussalam; Disease Control Division, Ministry of Health, Brunei Darussalam; Western Pacific Regional Office (Manila), World Health Organization

**Author notes:** Corresponding author: Dr Justin Wong, Disease Control Division, Ministry of Health, Commonwealth Drive, Bandar Seri Begawan BB3910, Brunei Darussalam.

## Abstract

**Background:** Current SARS-CoV-2 containment measures rely on the capacity to control person-to-person viral transmission. Effective prioritization of these measures can be determined by understanding SARS-CoV-2 transmission dynamics. We conducted a systematic review and meta-analyses of three parameters: (i) secondary attack rate (SAR) in various settings, (ii) clinical onset serial interval (SI), and (iii) the proportion of asymptomatic infection.

**Methods and Findings:** We searched PubMed, medRxiv, and bioRxiv databases between January 1, 2020, and May 15, 2020, for articles describing SARS-CoV-2 attack rate, SI, and asymptomatic infection. Studies were included if they presented original data for estimating point estimates and 95% confidence intervals of the three parameters. Random effects models were constructed to pool SAR, mean SI, and asymptomatic proportion. Risk ratios were used to examine differences in transmission risk by setting, type of contact, and symptom status of the index case. Publication and related bias were assessed by funnel plots and Egger’s meta-regression test for small-study effects.

Our search strategy for SAR, SI, and asymptomatic infection identified 459, 572, and 1624 studies respectively. Of these, 20 studies met the inclusion criteria for SAR, 18 studies for SI, and 66 studies for asymptomatic infection. We estimated the pooled household SAR at 15.4% (95% CI: 12.2%, 18.7%) compared to 4.0% (95% CI: 2.8%, 5.2%) in non-household settings. We observed variation across settings; however, the small number of studies limited power to detect associations and sources of heterogeneity. SAR of symptomatic index cases is significantly higher than cases that were symptom-free at diagnosis (RR 2.55, 95% CI: 1.47, 4.45). Adults appear to be more susceptible to transmission than children (RR 1.40, 95% CI: 1.00, 1.96). The pooled mean SI is estimated at 4.87 days (95% CI: 3.98, 5.77). The pooled proportion of cases who had no symptoms at diagnosis is 25.9% (95% CI: 18.8%, 33.1%).

**Conclusions:** Based our pooled estimates, 10 infected symptomatic persons living with 100 contacts would result in 15 additional cases in <5 days. To be effective, quarantine of contacts should occur within 3 days of symptom onset. If testing and tracing relies on symptoms, one-quarter of cases would be missed. As such, while aggressive contact tracing strategies may be appropriate early in an outbreak, as it progresses, control measures should transition to account for SAR variability across settings. Targeted strategies focusing on high-density enclosed settings may be effective without overly restricting social movement.

## Introduction

The ongoing pandemic caused by SARS-CoV-2 continues to escalate. Modeling studies have enhanced understanding of COVID-19 transmission dynamics and initial phylogenetic analysis of closely related viruses suggest highly linked person-to-person spread of SARS-CoV-2 originating from mid-November to early December 2019 (1-3).

There are no known effective therapeutics or vaccines (4, 5). As such, containment measures rely on the capacity to control viral transmission from person-to-person, such as case isolation, contact tracing and quarantine, and physical distancing (6). Effective prioritization of these measures can be determined by understanding SARS-CoV-2 transmission patterns.

There is an abundance of literature on the biological mode of transmission of coronaviruses: through exhaled droplets, aerosol at close proximity, fomites, and possibly through fecal-oral contamination (7, 8). However, few observational studies have assessed transmission patterns in populations, and what determines whether the infection is contained or spreads. Previous theoretical work by Fraser et al. proposed three transmission-related criteria that impact on outbreak control: (i) viral transmissibility; (ii) disease generation time; and (iii) the proportion of transmission occurring prior to symptoms (9).

To better understand SARS-CoV-2 transmission, we conducted a systematic review and meta-analyses of these three parameters. Using publicly available articles, we estimated the (i) secondary attack rate (SAR) in various settings, (ii) clinical onset serial interval (SI) from studies that linked dates of symptom onset for infector-infectee pairs, and (iii) proportion of asymptomatic infection.

## Methods

This systematic review and meta-analysis followed the Preferred Reporting Items for Systematic Reviews and Meta-Analyses (PRISMA) guidelines.

### Definitions

SAR is defined as the probability that an exposed susceptible person develops disease caused by an infected person (10). It is calculated by dividing the number of exposed close contacts who tested positive (numerator) by the total number of close contacts of the index case (denominator).

SI is defined as the time between disease symptom onset of a case and that of its infector (11). It is used as a proxy for the generation interval—the time between the infection of the infectee and that of the infector, which is an unobservable duration.

Asymptomatic cases are defined as those with laboratory confirmation, but without clinical signs and symptoms at diagnosis. This definition therefore includes pre-symptomatic cases. The asymptomatic proportion is calculated as the number of asymptomatic cases divided by the total number of cases.

### Data source and search strategy

We performed a literature search of published journal articles in PubMed and pre-print articles in medRxiv and bioRxiv from January 1, 2020. For SAR, we used the search terms (“SARS-CoV-2” OR “COVID-19”) AND (“attack rate” OR “contact tracing”). We replaced the last search term with “serial interval” for SI, and “asymptomatic” for asymptomatic infection. The last search date was on May 15, 2020. All studies that were written in English or have an abstract in English were included.

The articles were initially screened by title and abstract, and subsequently by review of selected full-text articles. Three reviewers selected the studies independently using predetermined inclusion criteria and differences in opinions were resolved through consensus. From the included articles, the same reviewers extracted the data for all parameter estimates: event counts, point estimates, confidence intervals (CI), and sample size. Whenever available, we also extracted data on setting, symptom status, age group, and relationship of cases and close contacts. Data were obtained directly from the reports, but when not explicitly stated, we derived the data from tables, charts, or supplementary materials.

### Inclusion criteria

Studies reporting SAR were included if they: (i) presented original data for estimating the SAR, such as from a contact tracing investigation; (ii) reported a numerator and denominator of close contacts, or at least two of numerator, denominator, and SAR; (iii) specified a particular setting; and (iv) cases were confirmed positive with SARS-CoV-2 through reverse transcription polymerase chain reaction (RT-PCR) test. Point-testing or prevalence studies to measure cumulative incidence of infection in a setting were excluded as the source of infection could not be traced, but we reported these studies as supplementary materials.

For SI, studies were included if they: (i) contained original parameter estimates of either mean or median SI; (ii) reported the associated 95% CI; and (iii) reported the number of infector-infectee pairs.

For asymptomatic infection, studies were included if they: (i) presented original data for estimating the proportion of asymptomatic cases at diagnosis, such as from a cross-sectional survey, cohort study, or case series; (ii) reported the number of asymptomatic cases and the number of total cases, or at least two of the asymptomatic proportion, number of asymptomatic cases, and number of total cases; and (iii) cases were confirmed positive by RT-PCR test. Studies with data on asymptomatic infection assessed at admission or throughout the follow-up period were included for comparison purposes.

### Statistical analysis

Point estimates and 95% CI were calculated. Meta analyses were performed using a random-effects DerSimonian-Laird model (12) for the SAR, SI, and the proportion of asymptomatic cases. We estimated risk ratios to examine SAR differences by setting, symptom status of the index case, age of close contacts, and relationship of household contacts. The I^2^ statistic is used as a measure of heterogeneity, with higher values signifying greater degree of variation (13). Publication and related bias were assessed by funnel plots and Egger’s meta-regression test for small-study effects (14). A p-value of <0.05 was considered as statistically significant. Statistical analysis was done in STATA 14 using the package metan, metafunnel, and metabias (15-17).

## Results

### Secondary attack rate

We identified 20 studies that allowed direct estimation of the SAR (Table 1; Supplementary materials Figure S1a). Fifteen of these were published articles (three in Chinese language) and the other five were pre-prints. Some studies identified close contacts through active surveillance systems while in others they were identified following an outbreak investigation. Testing protocols of close contacts also differed; in the United States, only those with symptoms or persons under investigation were tested whereas in others, particularly Asian countries, all close contacts were tested regardless of symptoms.

**Table 1.**
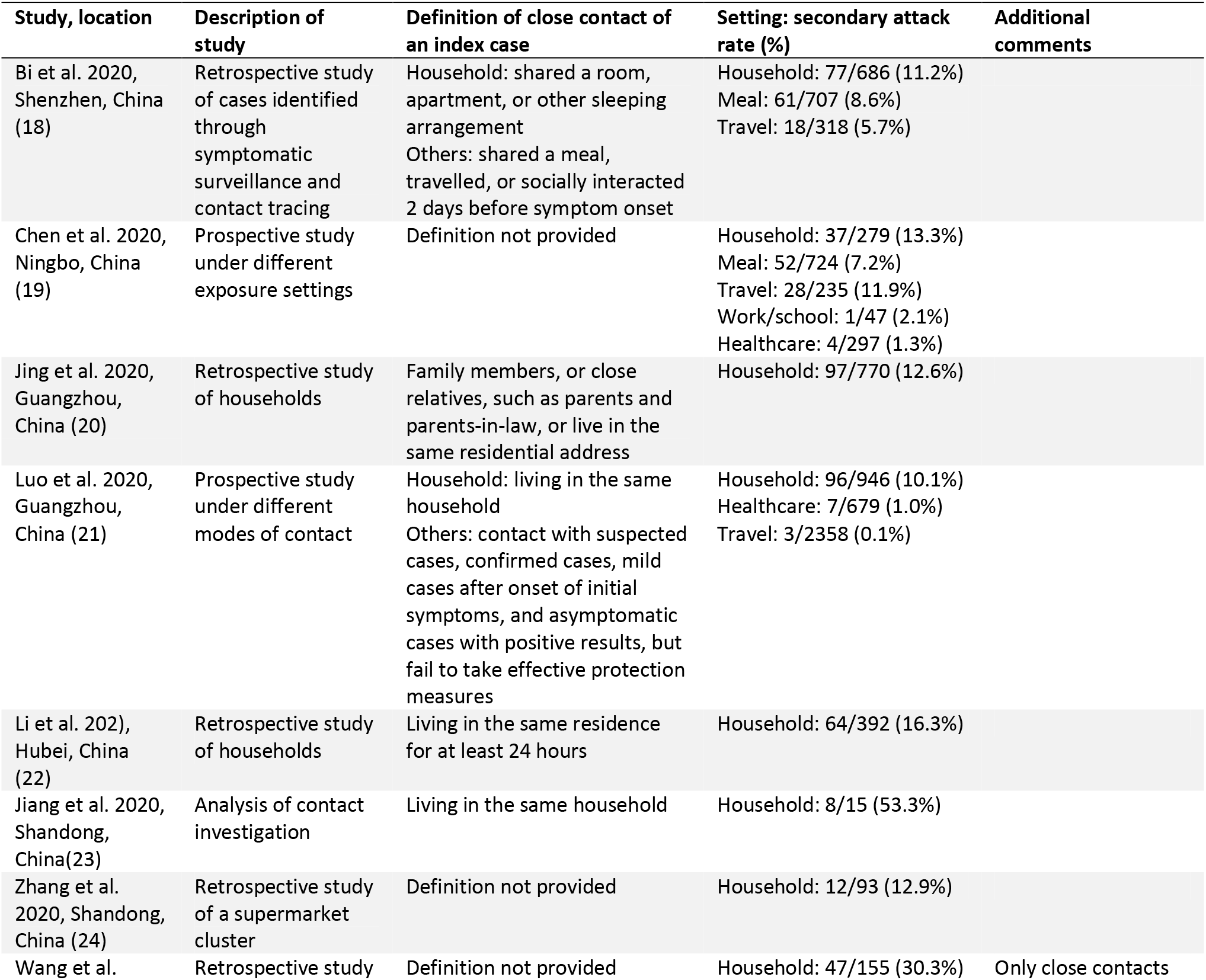
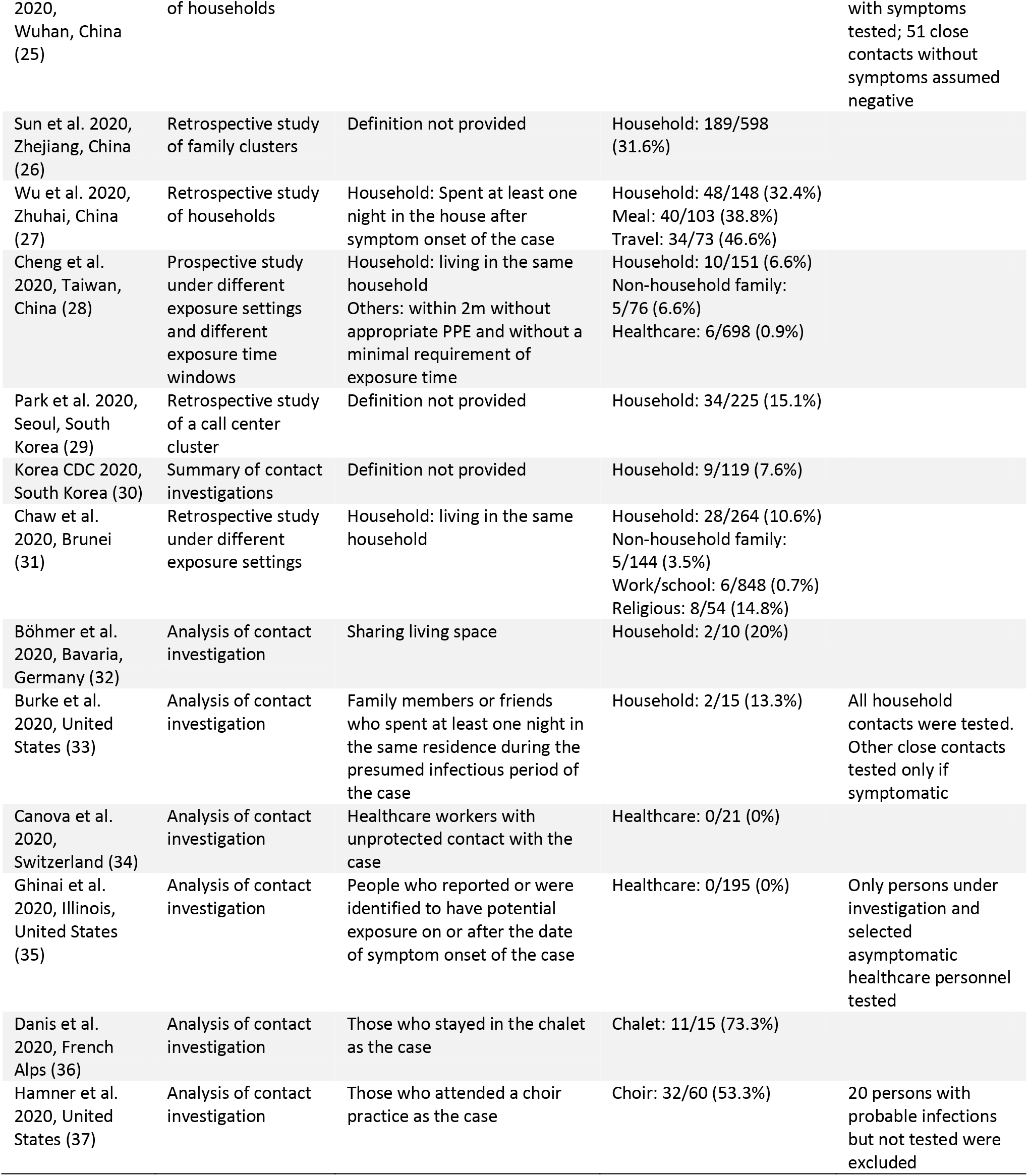
Description of studies included in the review and analysis of secondary attack rate (SAR)

There was variation in the definition of household contacts; most used a traditional definition as those who resided with the index case but some studies broadened the scope to include others who spent at least a night in the same residence or a specified duration of at least 24 hours of living together.

From these studies, we estimated household SAR in 17 locations, and non-household SAR in different settings in 10 locations. The non-household settings were non-household family, meal, travel, workplace or school, healthcare, religious event, chalet, and choir. The sample size of the settings varied depending on the location and context studied. We also estimated the SAR by symptom status of the index case, adults vs. children, and in the household, spouse vs. child and other household members.

Figure 1 summarizes the estimated SARs. The estimated mean household SAR is 15.4% (95% CI: 12.2%, 18.7%) with significant heterogeneity (p <0.001). Household SAR ranged from 6.6% in Taiwan to more than 30% in Shandong, Wuhan, Zhejiang, and Zhuhai in mainland China. In non-household settings, the mean SAR is significantly lower at 4.0% (95% CI: 2.8%, 5.2%). However, there is large variation across the settings. Higher SARs were observed in a chalet (73.3%), at a choir (53.3%), eating with a case (18.2% average), and at a religious event (14.8%). Funnel plot and Egger’s meta-regression test for household SAR studies do not support the presence of publication bias and small-study effects (Supplementary materials Figure S2a, Table S1a). Similar analyses could not be done for other settings due to the limited number of studies.

**Figure 1.**
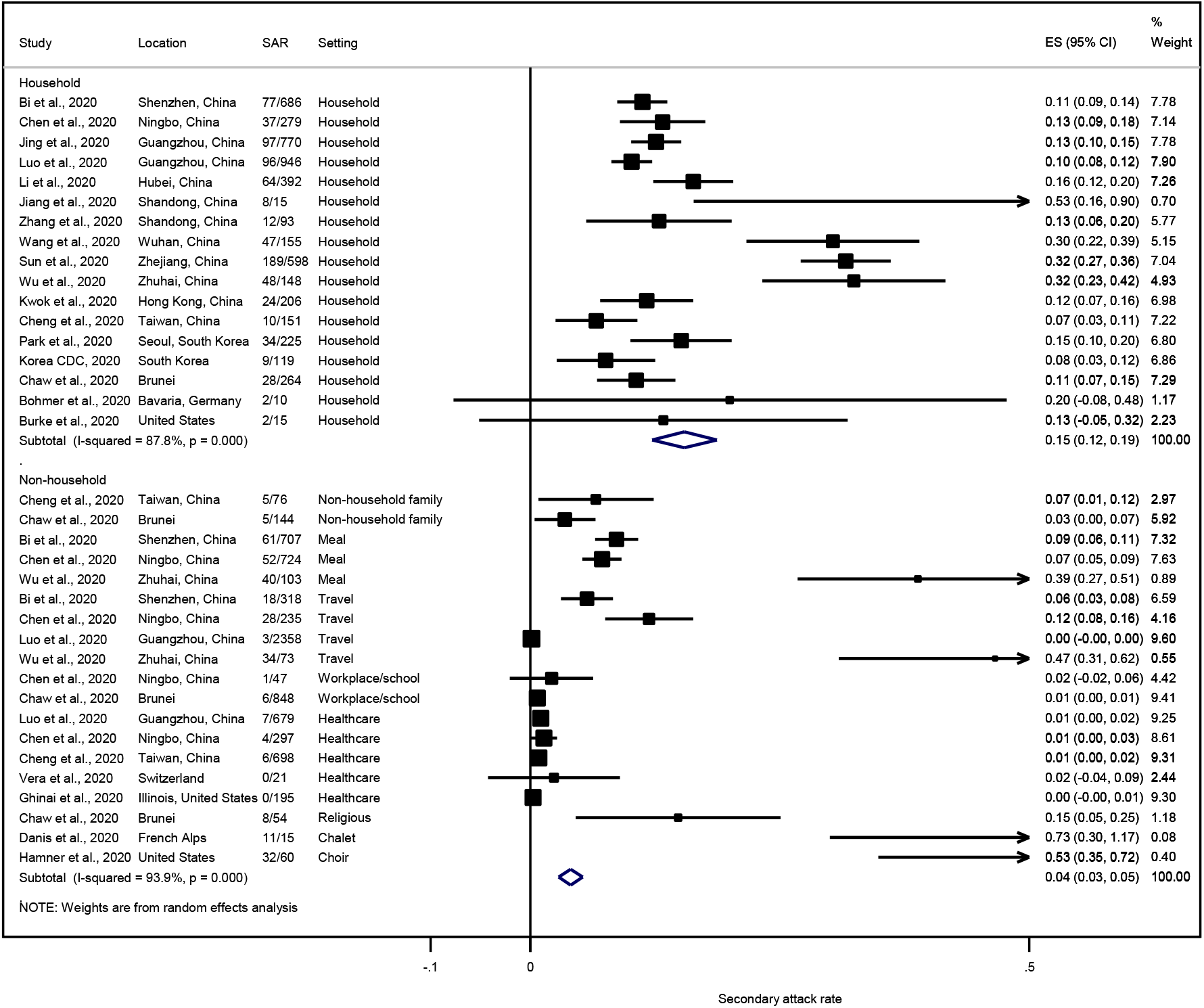
Forest plot of secondary attack rates (SAR) in different settings. ES is the estimated SAR, with 95% confidence intervals (CI). I-squared is the percentage of between-study heterogeneity that is attributable to variability in the true effect, rather than sampling variation.

The risk of infection also varies by the symptom status of the index case. Based on six studies, SARs of symptomatic index cases were 2.55 times of that of asymptomatic cases (including pre-symptomatic cases; Supplementary materials Figure S3).

SARs from eight studies showed that close contacts who are adults were more likely to be infected compared to children, with a relative risk of 1.40 (95% CI: 1.00, 1.96) (Supplementary materials Figure S4). SARs from several studies indicated a 3.23 (95% CI: 2.23, 4.68) times risk of infection for a spouse of an index case as compared to a child (Supplementary materials Figure S5), and 3.63 (95% CI: 1.68, 7.87) times as compared to other household members (Supplementary materials Figure S6).

### Serial interval

We included 18 studies (9 published articles, 9 pre-prints) in the meta-analysis of the mean SI (Table 2; Supplementary materials Figure S1b). We found a pre-print rapid review of SI (38) and cross-checked their articles—they were all included in our full-text shortlist. Several studies used the same dataset of the infector-infectee pairs; we therefore selected those that were the most comprehensive from a single location. For studies with multiple locations, publicly available datasets were used and hence there is likely to be substantial overlaps; nonetheless, we reported these studies as a comparison instead of discarding them.

**Table 2.**
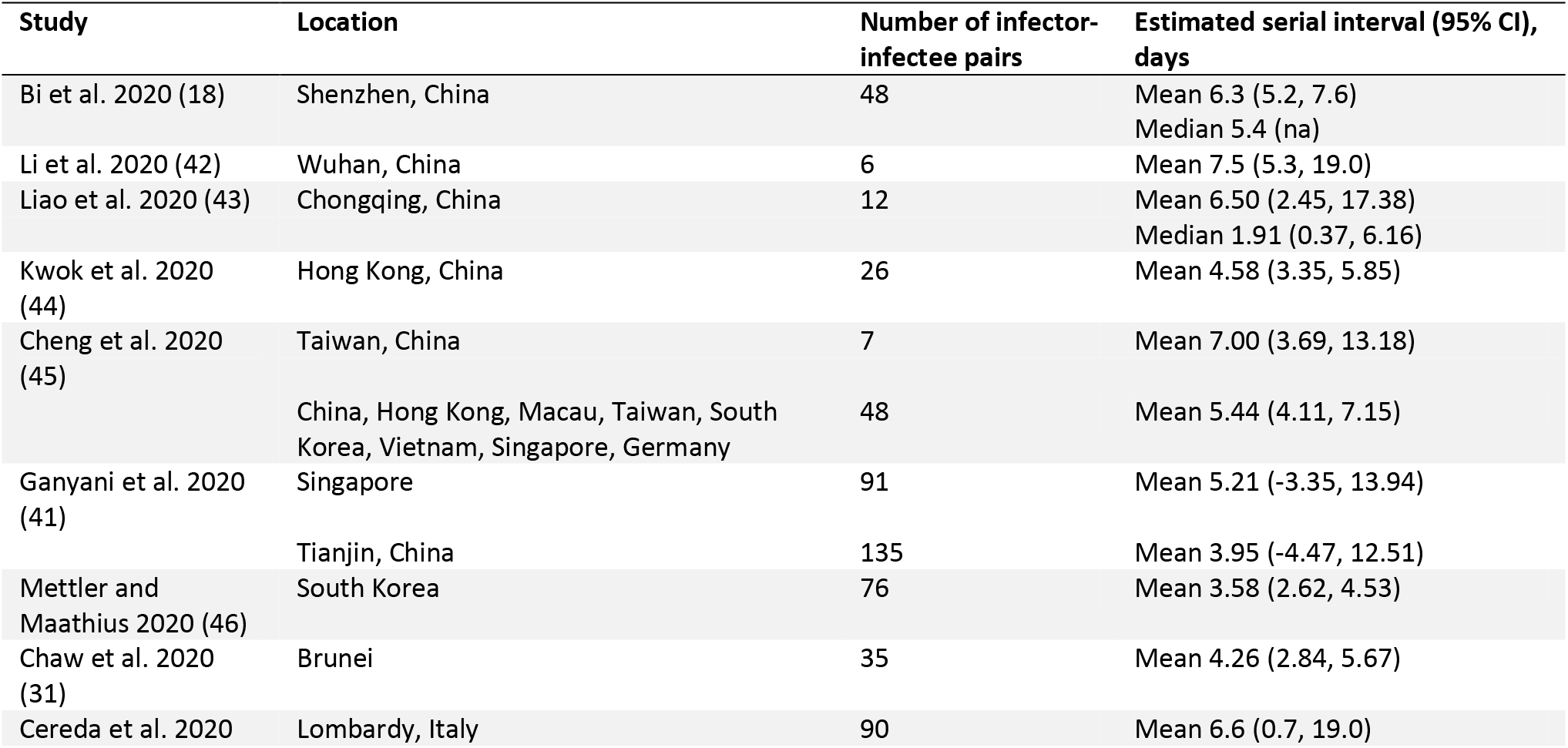
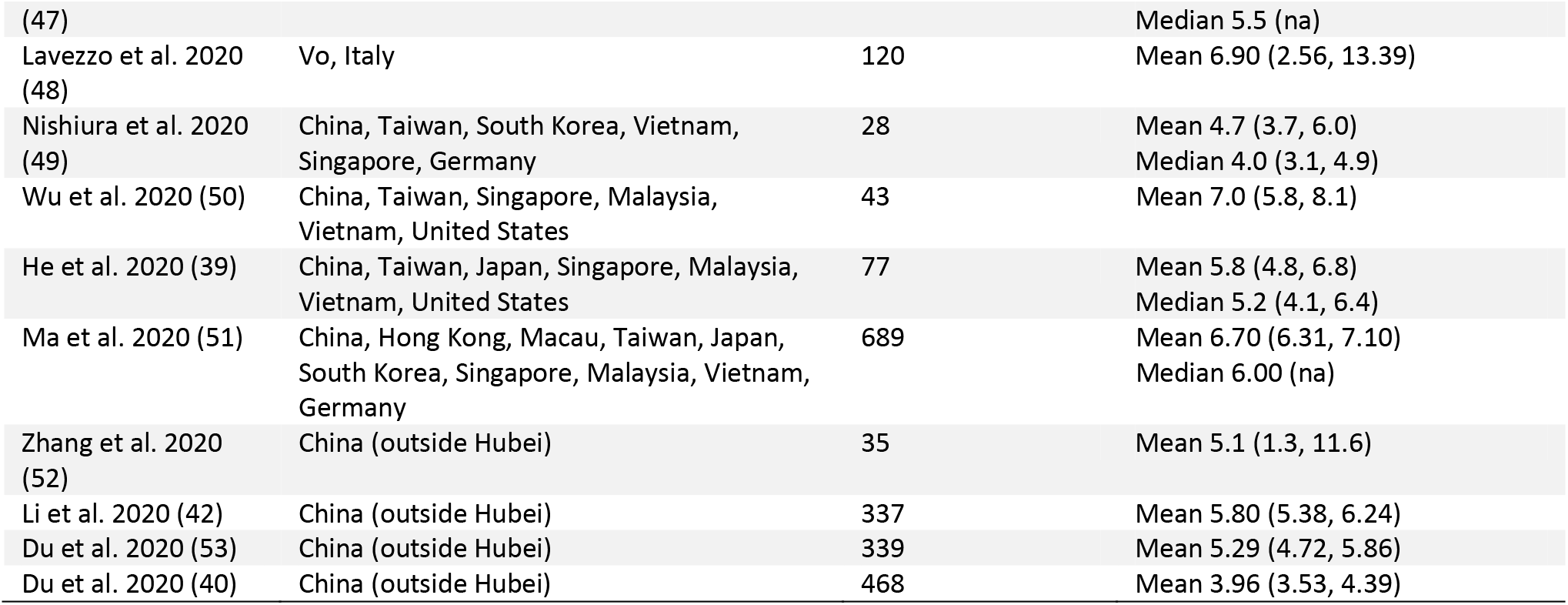
Description of studies included in the review and analysis of the serial interval (SI)

Most of the studies were from Asian countries, particularly China, and two were from Italy. In multiple-location studies, data from Germany and the United States were also used. The number of infector-infectee pairs ranged from less than 10 in single-location studies to 689 in multiple-location ones. However, studies with a large sample size are not necessarily representative of the countries from which the data were drawn. For instance, in He et al. with 77 pairs used, only one was from the United States, 20 were from East Asian countries, and the rest from mainland China (39). We evaluated the value of a larger sample size against the accuracy of the infector-infectee relationship, and decided to retain all small-sample studies.

The mean SI was more often reported compared to the median. Statistical distributions used were usually Gamma, Weibull, or lognormal. Negative serial intervals were reported in a few studies, and the authors suggested the Normal distribution as a more appropriate fit (31, 40, 41).

The estimated mean SI are shown in Figure 2. The mean SI of the single-location studies is estimated to be 4.87 days (95% CI: 3.98, 5.77). The shortest SI was observed in South Korea (3.6 days) and the longest in Wuhan (7.5 days). In the multiple-location studies, the shortest mean SI was 4.0 days and longest 7.0 days. There is wide heterogeneity in the multiple-location studies (p <0.001), but less in the single-location ones.

**Figure 2.**
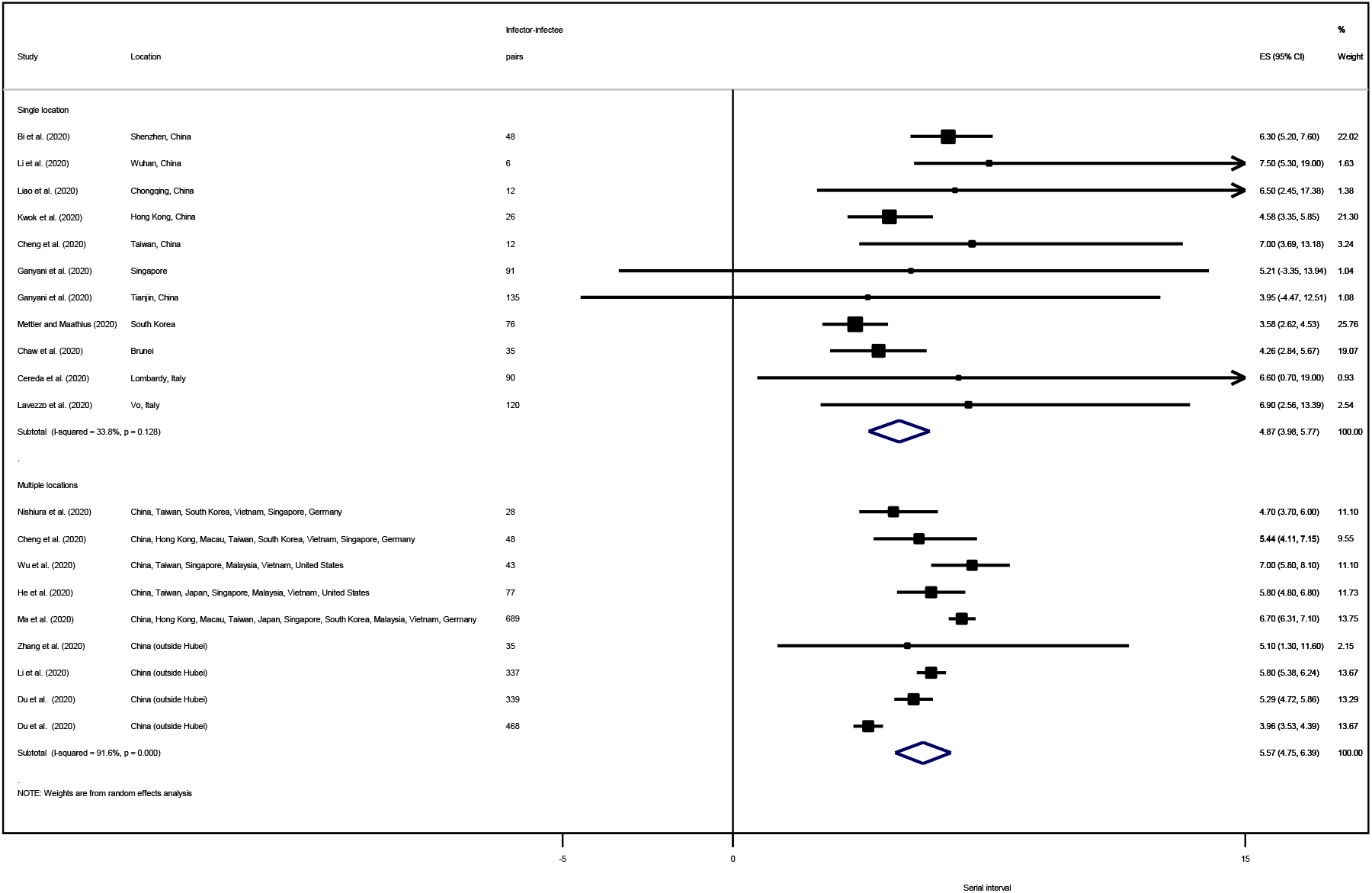
Forest plot of the serial interval (SI) ES is the estimated mean SI, with 95% confidence intervals (CI). I-squared is the percentage of between-study heterogeneity that is attributable to variability in the true effect, rather than sampling variation.

### Asymptomatic infection

We identified 66 studies that allowed estimation of the proportion of cases that were asymptomatic (Table 3; Supplementary materials Figure S1c). 54 of these were published articles (5 in Chinese language) and the remainder pre-prints. Twelve studies focused on children. For studies with overlapping datasets, we selected the ones that were most comprehensive. We found a rapid review of asymptomatic infections (54) but it only included studies with cases that were asymptomatic throughout the duration of their illness; we cross-checked the articles and they were all covered in our full-text shortlist.

**Table 3.** Description of studies included in the review and analysis of asymptomatic infection.

| Study | Location | Type of study | Study population | Mean/median age of study population (range) | Asymptomatic proportion (%) | Time of symptom assessment |
| --- | --- | --- | --- | --- | --- | --- |
| Bai et al. 2020 (55) | Anyang, China | Retrospective study | Family cluster | na | 1/6 (16.7%) | Diagnosis |
| Luo et al. 2020 (56) | Anhui, China | Retrospective study | Family cluster | na | 2/4 (50%) | Diagnosis |
| Zhang et al. 2020 (57) | Beijing, China | Retrospective study | Family cluster | na | 2/5 (40%) | Diagnosis |
| Bai et al. 2020 (58) | Gansu, China | Retrospective study | Family cluster | 47 (2-82) | 5/8 (62.5%) | Diagnosis |
| Pan et al. 2020 (59) | Guangzhou, China | Retrospective study | Family cluster | 23.7 (3-35) | 2/3 (66.7%) | Diagnosis |
| Jiang et al. 2020 (60) | Henan, China | Retrospective study | Family cluster | 48 (9-87) | 4/9 (44.4%) | Diagnosis |
| Ye et al. 2020 (61) | Luzhou, China | Retrospective study | Family cluster | 40.4 (23-51) | 3/5 (60%) | Diagnosis |
| Jiang et al. 2020 (23) | Shandong, China | Retrospective study | Family cluster | 39.4 (0.25-65) | 5/8 (62.5%) | Diagnosis |
| Chan et al. 2020 (62) | Shenzhen, China | Retrospective study | Family cluster | 48 (10-66) | 1/5 (20%) | Admission |
| Luo et al. 2020 (63) | Wuhan, China | Retrospective study | Family cluster | 36 (7-64) | 5/6 (83.3%) | Diagnosis |
| Qian et al. 2020 (64) | Zhejiang, China | Retrospective study | Family cluster | 46.6 (1-76) | 2/8 (25%) | Diagnosis |
| Park et al. 2020 (29) | Seoul, South Korea | Contact tracing | Employees at a call center | 38 (20-80) | 8/97 (8.2%) | Diagnosis |
| Danis et al. 2020 (36) | French Alps | Contact tracing | Guests in a chalet | na | 1/12 (8.3%) | Diagnosis |
| Yamahata et al. 2020 (65) | Yokohama, Japan | Point-testing | Cruise passengers (Diamond Princess) | na | 410/696 (58.9%) | Diagnosis |
| Lombardi et al. 2020 (66) | Lombardy, Italy | Point-testing | Healthcare workers | 44.5 (21-76) | 41/138 (29.7%) | Diagnosis |
| Arons et al. 2020 (67) | King County, Washington | Point-testing | Residents in a nursing facility | 78.6 (na) | 27/48 (56.3%) | Diagnosis |
| Roxby et al. 2020 (68) | Seattle, Washington | Point-testing | Residents in an independent and assisted living community | 68.3 (51-92) | 3/6 (50%) | Diagnosis |
| Hamner et al. 2020 (37) | Skagit County, Washington | Contact tracing | Members of a choir practice | 69 (31-83) | 8/61 (13.1%) | Throughout |
| Arima et al. 2020 (69) | Japan | Point-testing | Residents evacuated from China | na | 5/7 (71.4%) | Diagnosis |
| Zhang et al. 2020 (24) | Liaocheng City, China | Contact tracing | Supermarket cluster | na | 3/25 (12%) | Diagnosis |
| Ly et al. 2020 (70) | Marseille, France | Cross-sectional survey | Vulnerable population | na | 25/49 (51%) | Diagnosis |
| Breslin et al. 2020 (71) | New York | Retrospective study | Pregnant patients from New York City hospitals | 26.9 (20-39) | 14/43 (32.6%) | Admission |
| Wu et al. 2020 (72) | Wuhan, China | Retrospective study | Pregnant patients from the Maternal and Child Health Hospital of Hubei Province | 29.9 (26-35) | 4/8 (50%) | Admission |
| Wu et al. 2020 (73) | Wuhan, China | Retrospective study | Pregnant patients from the Central Hospital of Wuhan | 29 (21-37) | 15/23 (65.2%) | Admission |
| Huang et al. 2020 (74) | Anhui, China | Contact tracing | Cases and contacts in Anhui | 22 (16-23) | 0/8 (0%) | Admission |
| Tian et al. 2020 (75) | Beijing, China | Retrospective study | Patients from Beijing hospitals | 47.5 (1-94) | 13/262 (5%) | Throughout |
| Chen et al. 2020 (76) | Chongqing, China | Retrospective study | Patients from Chongqing hospitals | 47 (37-61) | 30/136 (22.1%) | Admission |
| Liao et al. 2020 (43) | Chongqing, China | Retrospective study | Patients from Chongqing Three Gorges Central Hospital of Chongqing University | na (10-35) | 4/46 (8.7%) | Admission |
| Chen et al. 2020 (77) | Guangzhou, China | Retrospective study | Patients from Guangzhou Eighth People's Hospital | 48 (na) | 17/284 (6%) | Throughout |
| Luo et al. 2020 (21) | Guangzhou, China | Prospective study | Cases and close contacts in Guangzhou | na | 8/129 (6.2%) | Throughout |
| Kim et al. 2020 (78) | Gwangju, South Korea | Retrospective study | Patients from Affiliated Hospitals of Chonnam National University | na | 13/71 (18.3%) | Diagnosis |
| Li et al. 2020 (22) | Hubei, China | Contact tracing | Household contacts in Hubei | 45 (na) | 9/64 (14.1%) | Throughout |
| Wan et al. 2020 (79) | Hunan, China | Retrospective study | Patients from the First Affiliate Hospital of Hunan Normal University | na | 2/78 (2.6%) | Throughout |
| Qiu et al. 2020 (80) | Hunan, China | Contact tracing | Patients from the First People's Hospital of Huaihua and the Central Hospital of Shaoyang | 43 (8-84) | 5/104 (4.8%) | Throughout |
| Yin et al. 2020 (81) | Hunan, China | Retrospective study | Patients from Second Xiangya Hospital of Central South University | 46 (31.5-65) | 4/33 (12.1%) | Throughout |
| Xu et al. 2020 (82) | Jiangsu, China | Retrospective study | Patients from Jiangsu hospitals | na | 15/342 (4.4%) | Throughout |
| Ma et al. 2020 (83) | Jinan, China | Retrospective study | Patients from Jinan Infectious Diseases Hospital | 34 (1-72) | 11/47 (23.4%) | Throughout |
| Yin and Jin 2020 (84) | Ningbo, China | Contact tracing | Cases and close contacts in Ningbo | na | 30/187 (16%) | Diagnosis |
| Du et al. 2020 (85) | Shandong, China | Retrospective study | Patients from Jinan Infectious Diseases Hospital and Rizhao People's Hospital | 34.1 (na) | 8/67 (11.9%) | Throughout |
| Zhou et al. 2020 (86) | Shanghai, China | Retrospective study | Patients from Shanghai Public Health Center | na | 13/328 (4%) | Diagnosis |
| Bi et al. 2020 (18) | Shenzhen, China | Contact tracing | Cases and close contacts in Shenzhen | 45 (na) | 25/391 (6.4%) | Admission |
| Song et al. 2020 (87) | Tibetan Autonomous Prefecture, China | Retrospective study | Patients from People's Hospital in Daofu county | na | 18/83 (21.7%) | Throughout |
| He et al. 2020 (88) | Wenzhou, China | Retrospective study | Patients from hospitals in Wenzhou | na | 12/206 (5.8%) | Diagnosis |
| Tao et al. 2020 (89) | Wuhan, China | Retrospective study | Patients from Chongqing Public Health Medical Center | 46 (na) | 20/167 (12%) | Throughout |
| Wang et al. 2020 (90) | Wuhan, China | Retrospective study | Patients from Fangcang Hospital | 50 (16-89) | 30/1012 (3%) | Throughout |
| Xu et al. 2020 (91) | Wuhan, China | Retrospective study | Patients from hospitals in several provinces | 57 (43-69) | 1/69 (1.4%) | Admission |
| Zhang et al. 2020 (92) | Wuhan, China | Retrospective study | Patients from Renmin Hospital of Wuhan University | 45.4 (na) | 16/120 (13.3%) | Admission |
| Wu et al. 2020 (27) | Zhuhai City, China | Contact tracing | Cases and contacts in Zhuhai | 45.8 (na) | 8/83 (9.6%) | Throughout |
| Leung et al. 2020 (93) | Hong Kong, China | Retrospective study | Patients from Hong Kong hospitals | 55.2 (22-96) | 2/50 (4%) | Throughout |
| Liu et al. 2020 (94) | Taiwan | Case series | Imported cases in Taiwan | na (4-88) | 11/321 (3.4%) | Throughout |
| Korea CDC 2020 (95) | South Korea | Case series | Confirmed cases in South Korea | 42.6 (20-73) | 3/28 (10.7%) | Throughout |
| Kim et al. 2020 (96) | Daegu, South Korea | Cross-sectional survey | Patients at an isolation facility | 26 (na) | 41/213 (19.2%) | Admission |
| Qasim et al. 2020 (97) | Japan | Case series | Confirmed cases in Japan | na (40-79) | 112/1192 (9.4%) | Diagnosis |
| Wong et al. 2020 (98) | Brunei | Case series | Confirmed cases in Brunei | na | 58/138 (42%) | Diagnosis |
| Pongpirul et al. 2020 (99) | Thailand | Retrospective study | Patients from Bamrasnaradura Infectious Diseases Institute | 61 (28-74) | 1/11 (9.1%) | Throughout |
| Le et al. 2020 (100) | Vietnam | Retrospective study | Patients from hospitals in Vietnam | 31.2 (0.25-55) | 1/12 (8.3%) | Diagnosis |
| Shabrawishi et al. 2020 (101) | Saudi Arabia | Retrospective study | Patients from Al-Noor Specialist Hospital | 46.1 (11-87) | 47/150 (31.3%) | Throughout |
| Tan et al. 2020 (102) | Changsha, China | Retrospective study | Children (<12 years old) | 7 (1-12) | 2/10 (20%) | Throughout |
| Liao et al. 2020 (43) | Chongqing, China | Contact tracing | Children (<24 years old) | na (10-24) | 2/14 (14.3%) | Admission |
| Song et al. 2020 (103) | Hubei, China | Retrospective study | Children (<14 years old) | 8.5 (1-14) | 8/16 (50%) | Throughout |
| Du et al. 2020 (85) | Shandong, China | Retrospective study | Children (<16 years old) | 6.2 (0-16) | 8/14 (57.1%) | Throughout |
| Feng et al.<br>2020 (104) | Shenzhen,<br>China | Retrospective<br>study | Children (<14 years old) | 7.9 (4-14) | 10/15 (66.7%) | Diagnosis |
| Zhou et al.<br>2020 (105) | Shenzhen,<br>China | Retrospective<br>study | Children (<3 years old) | 1.6 (0.75-3) | 5/9 (55.6%) | Throughout |
| Lu et al.<br>2020 (106) | Wuhan,<br>China | Retrospective<br>study | Children (<15 years old) | 6.7 (0-15) | 27/171<br>(15.8%) | Diagnosis |
| Ma et al.<br>2020 (107) | Wuhan,<br>China | Retrospective<br>study | Children (<14 years old) | na (0-15) | 61/115 (53%) | Diagnosis |
| Tao et al.<br>2020 (89) | Wuhan,<br>China | Retrospective<br>study | Children (<14 years old) | na | 2/7 (28.6%) | Throughout |
| Xia et al.<br>2020 (108) | Wuhan,<br>China | Retrospective<br>study | Children (<14 years old) | 2.1 (0-14.5) | 2/20 (10%) | Throughout |
| Qiu et al.<br>2020 (109) | Zhejiang,<br>China | Retrospective<br>study | Children (<16 years old) | 8.3 (1-16) | 10/36 (27.8%) | Admission |
| Li et al.<br>2020 (110) | Zhuhai City,<br>China | Retrospective<br>study | Children (<6 years old) | 3.4 (1-6) | 4/5 (80%) | Admission |

Most of the retrospective cohort studies were from China and those on point-testing and prevalence studies were outside China. In terms of symptom assessment, many of the retrospective studies followed up patients throughout whereas symptoms at diagnosis could be observed in cross-sectional surveys and in investigation of family clusters. There is wide variation in the sample size, from less than 10 in family clusters in China to 1,192 in Japan’s case series. We did not exclude studies based on an arbitrary sample size cut-off as we see value in assessing asymptomatic infection in different settings and study populations.

Figure 3 summarizes the proportion of asymptomatic cases. The estimated mean asymptomatic proportion at diagnosis is 25.9% (95% CI: 18.8%, 33.1%). Unsurprisingly, this proportion is larger than that at admission (14.8%) and throughout the follow-up period (8.0%). In general, the asymptomatic proportion decreases with age—this is more evident in hospital-based studies that observe patients over a certain time period (Supplementary materials Figure S7). The mean asymptomatic proportion at diagnosis is higher in children at 41.8% but there is wide variation due to the limited number of studies (Supplementary materials Figure S8). The asymptomatic proportion in children seems to change little over the course of the follow-up period.

Among the studies with asymptomatic cases assessed at diagnosis, the asymptomatic proportion is notably high in the Diamond Princess cruise (58.9%), a nursing facility in the United States (56.3%), vulnerable population in France (51.0%), Japanese residents evacuated from China (33.3%), and healthcare workers in Italy (29.7%). Pregnant women also had a high asymptomatic proportion (49.3%). The proportion is also high in studies investigating family clusters (49.1%); we note that there is a systematic overestimation in such contact investigations since all clusters include at least one asymptomatic case.

The funnel plot for overall asymptomatic infection is asymmetric, suggesting the presence of small-study effects and confirmed by Egger’s meta-regression test (including segregation by asymptomatic at diagnosis, admission, or throughout), as noted above (Supplementary materials Figure S2b and Table S1b). We therefore conducted an additional search but could not identify any other relevant articles. There is no evidence of bias in studies on children (Supplementary materials Figure S2c and Table S1c).

**Figure 3.**
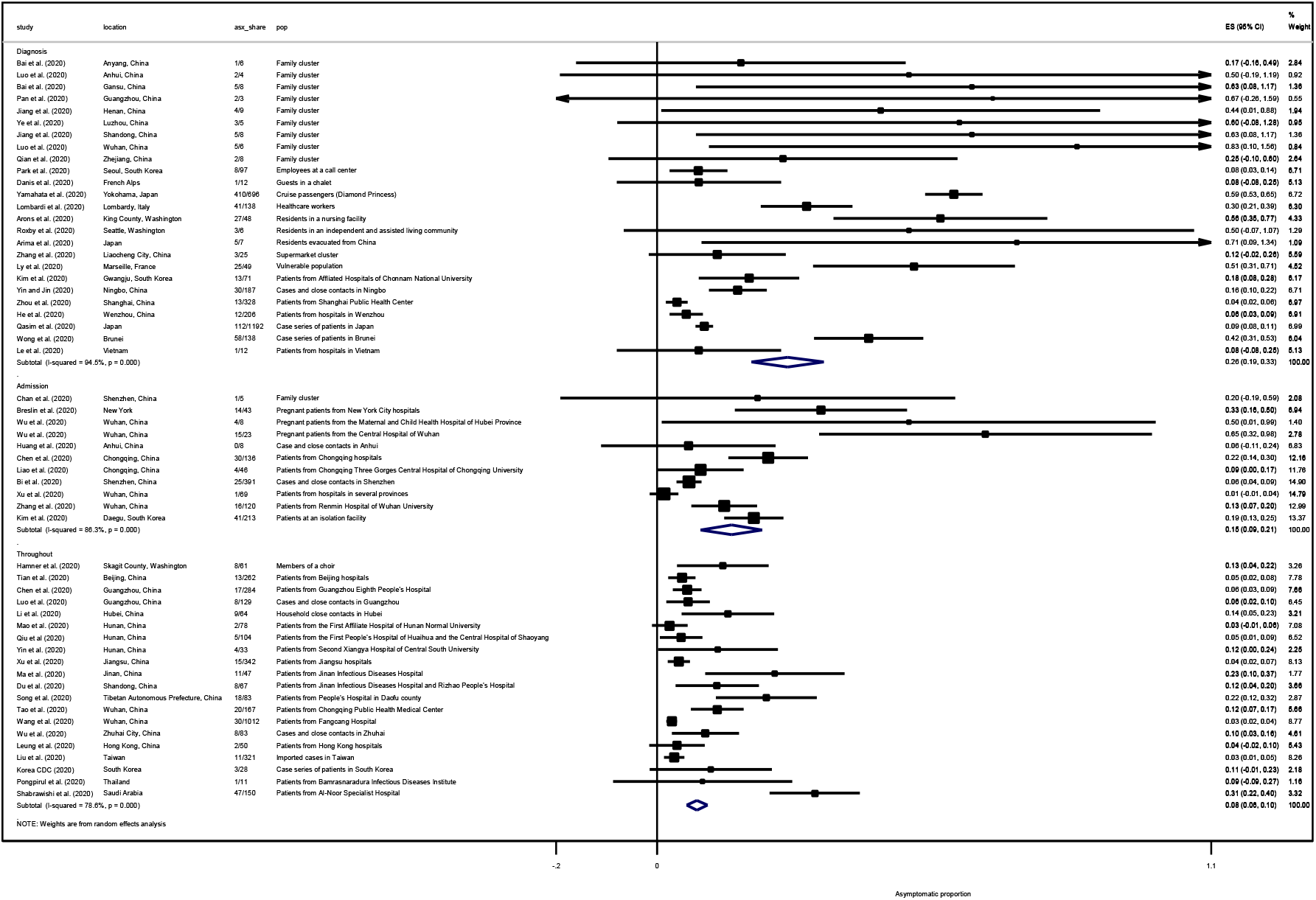
Forest plot of the proportion of asymptomatic cases. ES is the estimated asymptomatic proportion, with 95% confidence intervals (CI). I-squared is the percentage of between-study heterogeneity that is attributable to variability in the true effect, rather than sampling variation.

## Discussion

### Summary of key findings

We estimated household SAR at 15.4% (95% CI: 12.2%, 18.7%), almost four times higher than non-household SAR at 4.0% (95% CI: 2.8%, 5.2%). The mean SI was 4.87 days (95% CI: 3.98, 5.77) and the asymptomatic proportion at diagnosis was 25.9% (95% CI: 18.8%, 33.1%). Symptomatic persons had a 2.55 (95% 1.47, 4.45) times higher risk of infecting others.

### Secondary attack rate

We estimated SAR across various settings as a measure of viral transmissibility. While a number of studies have estimated the basic reproductive number (R0) at 2–4, (111-114) in isolation it is a suboptimal gauge of infectious disease dynamics as it does not account for variability in specific situations and settings (115, 116).

The significant heterogeneity in SAR across the different settings is unsurprising given that SAR depends not only on the causative agent but also on socio-demographic, environmental, and behavioral factors in study populations (117). Variation in methods for case ascertainment and then subsequent detection of infected cases among contacts likely contributed to the heterogeneity across studies.

Household SAR at 15.4% was higher than in non-household settings at 4.0%. Reports suggest that familial transmission account for the majority of transmissions (25, 118). The household is thought to be a fundamental unit of SARS-CoV-2 transmission because of the high frequency and intensity of contacts that occur between family members, and because transmission has continued in places with movement restriction (31). We found the household SAR for COVID-19 approaches the upper range of estimates of the household SAR for the 2009 H1N1 pandemic influenza (5–15%) (119-121), and higher than that observed for both SARS (5–10%) (122-124) and MERS (4–5%) (125, 126). This suggests relatively higher SARS-CoV-2 transmissibility in the household setting. SARS-CoV-2 also has a higher R0 when compared to MERS-CoV and SARS-CoV-1 (127). This finding highlights the necessity of case isolation, immediate tracing, and quarantine of household contacts (128).

The relatively lower SAR in non-household settings may mask variation between setting types. Some studies reported significantly higher SAR in mass gatherings and other enclosed settings with potential for prolonged physical contact, such as at the ski chalet in France (73.3%), at a choir in the United States (53.3%), and a religious gathering in Brunei (14.8%) (31, 36, 37). In contrast, SAR in workplace/school and in healthcare settings ranged between 1–2%, suggesting a gradation of risk outside the household (19, 28, 31). As an aggregate, we found that the transmission risk was significantly reduced outside the household.

Our meta-analysis excluded studies that solely reported attack rates (AR) without identification of an index case and their transmission generations within the cluster. However, such studies may be important in understanding the role of super-spreading events (SSEs) in driving SARS-CoV-2 transmission (116). Specific settings where high ARs have been observed were in a nursing home in Kings County, Washington (64%), a call centre in South Korea (43.5%), a church in Arkansas (38%), a homeless shelter in Boston (36%), a fitness dance class (26.3%), and the Diamond Princess cruise ship in Japan (18.8%) (Supplementary materials Table S2).

Reflecting on the high household SAR and high AR in some settings, we suggest that several common environmental factors potentially account for the rapid person-to-person transmission observed: closed environments, population density, and shared eating environments. This is supported by environmental sampling studies (129) and ecological observations on the declining incidence of COVID-19 cases in areas with restrictions on indoor mass gatherings (130).

There are implications for mass gatherings and certain religious events, particularly as countries begin to relax physical distancing measures. Non-household residential settings such as long-term care facilities, dormitories, and detention facilities pose specific challenges where additional prevention measures merit consideration, including staff screening, enhanced testing, and strict visitor policies (67).

Certainly, across all settings, the longer the duration and the greater the degree of physical contact with an index case, the higher the risk of transmission. However, we find that the risk model for transmission of SARS-CoV-2 is nuanced—while the highest risk of transmission is in crowded and enclosed settings, social interaction in public settings has a lower risk. This suggests that socially disruptive large-scale community lockdowns can be avoided if case isolation, tracing, and physical distancing measures that target higher risk activities can be implemented effectively (131). This combination of measures is dependent upon the stage of the epidemic in a particular area and may be more feasible in areas with imported cases or limited local transmission.

In addition, as anxiety with physical distancing measures (so-called “quarantine fatigue”) gain momentum (132), public communications surrounding these measures should convey this continuum of risk based on the transmission dynamics across different settings, supporting sustainable longer-term behavior changes.

### Serial interval

The SI depends upon the temporal relation between infectiousness and clinical onset of a source case and the incubation period of the receiving case (133). We estimated the mean SI at 4.87 days, with studies reporting the SI ranging from 3.58 to 7.5 days, which is considerably shorter than observed for SARS (8.4 days) and MERS (14.6 days). The variation between studies is expected, as the interval between symptoms in an infector-infectee pair is influenced by the level of close contact, varying widely between different countries and in different settings.

Negative serial intervals were reported in several studies indicating that the infectee had symptoms earlier than the infector, thereby strengthening the evidence for pre-symptomatic transmission (98). Using a mean SI of 5.21 days obtained from the Grace Assembly of God cluster in Singapore, Ganyani et al. conducted a sensitivity analysis suggesting that the proportion of pre-symptomatic transmission increased from 48% to 66% when negative SI values were taken into account (41). Other studies have estimated that persons can potentially be transmitting up to 2 days prior to symptom onset (28, 39). Taken together, our findings suggest that for quarantine to be effective, cases have to be identified and contacts traced and quarantined within 2.87 days of the onset of symptoms of the index case. Quarantine after this period potentially results in transmission from secondary to tertiary cases.

### Asymptomatic infections

If significant transmission were occurring prior to symptoms, this could pose challenges for symptom-based public health control measures, such as clinical criteria for testing, and early case isolation (9). We estimated that 25.9% of COVID-19 cases were asymptomatic at time of diagnosis. Of these, more than two-thirds went on to develop symptoms over the course of their disease with a ‘true asymptomatic’ proportion of 8.4%. Our study does not support claims that the majority of SARS-CoV-2 infections are asymptomatic (134). Questions remain as to the role of asymptomatic carriers in the transmission of SARS-CoV-2. A number of studies have described familial clusters resulting from asymptomatic carrier transmission (55, 135). From the observational studies included in our analysis, we found a significantly higher risk of transmission where the index case was symptomatic with a relative risk of 2.55, suggesting that testing strategies should prioritize symptomatic persons (9) particularly where resources are finite.

Nonetheless, the proportion of asymptomatic infection combined with a relatively short SI warrant caution as there is potential for silent transmission (98). Even with a highly effective case isolation and quarantine system, asymptomatic infections are difficult to detect rapidly and may give rise to transmission chains within community settings. Therefore, some degree of physical distancing is likely to be needed to account for this (131).

### SARS-CoV-2 transmission in children

For many infectious diseases, such as seasonal and pandemic influenza, children are known be drivers of transmission in households and communities (136). Case series data on SARS-CoV-2 suggests that children are less likely to be affected than adults. A national analysis of the first 72,314 cases in China reported only 2.1% of all cases were in children aged 0-19 years old (137). Other studies show similarly low proportions (138).

To better understand their relative susceptibility of infection, we compared the SAR between adults and children and found adults at 1.40 times higher risk of infection than children. The lower rate of susceptibility in children could be explained by differences in symptomatic infection rates and subsequent issues with case ascertainment (139). The estimated mean asymptomatic proportion among children is 41.8%, about 1.6 times the proportion in the general population.

The literature surrounding infectivity in children is scarce. In household transmission studies, children are usually identified through contact tracing of adult cases, although a number of case reports have documented transmission from children to adults (140). There is also insufficient knowledge on transmissibility of SARS-CoV-2 from children to other children. In addition, age may be important to determine dynamics of interactions among children but inadequate data hampered our efforts at risk stratification by age.

While there are still important unknowns with respect to SARS-CoV-2 in children, these early findings may assist health authorities in determining proportionate thresholds for school closures in future waves of the pandemic.

### Strengths and limitations

Our analysis has important limitations. The studies selected were based on field investigation; variability was noted with respect to study design, the number of individuals assessed, clinical definitions, the extent to which confirmatory laboratory tests were used, the methods of clinical data collection, and the duration of follow-up. Studies may have different definitions of household and contacts and are subject to recall and observer bias (141). Moreover, without accounting for outside sources of infection, setting-specific SARs are likely to be overestimated (117). In fact, none of the reviewed studies addressed the composition of secondary vs. community infections when estimating the SAR or used viral sequencing to confirm homology between the strains infecting the index and secondary cases in the household.

All studies on SAR were retrospective transmission studies based on contact tracing datasets where the index case determination or the direction of transmission may be uncertain, particularly in diseases such as COVID-19 where a substantial proportion of cases are asymptomatic or mild. An additional challenge concerns the timing of recruitment of cases and their contacts during the course of an epidemic. Studies conducted in early stages can provide timely SAR estimates; however, this may be influenced by behavioral factors and other non-pharmaceutical interventions (e.g. community quarantine) that alter over the course of the epidemic (117).

Studies reporting SI face have similar limitations. Most do not account for incomplete reporting over the course of the epidemic and hence have incomplete transmission networks (41). Changes in social contacts and other behavioral changes, such as mask-wearing, may also alter the SI (133). Akin to contact tracing-based transmission studies, the order of transmission in clusters can easily be mistaken.

Moreover, given the potential for pre-symptomatic and asymptomatic transmission, it can be difficult to determine the source of infection with certainty.

Estimates of the asymptomatic proportion should be interpreted with caution as they are approximated from studies that lack a standardized case definition for asymptomatic infections. The collection of acute and convalescent serology from household contacts in household transmission studies could provide key information on asymptomatic cases, which is essential to forecasting the course of the pandemic. We cannot exclude publication bias for studies reporting on asymptomatic proportion given the media attention surrounding this particular topic.

The major strength of our study is that it comprehensively covers several key parameters of SARS-CoV-2 transmission-related dynamics, thus facilitating discussion and allowing for triangulation of these different parameters to identify the key drivers of transmission.

### Conclusion

Our estimates of SAR, SI, and asymptomatic infection demonstrate the challenges in controlling SARS-CoV-2 transmission. Based on our pooled estimates of these three parameters, 10 infected symptomatic persons living with 100 contacts would result in 15 additional symptomatic infected persons in <5 days and 39 in <15 days. If the testing and tracing strategy is based on symptoms, we would further miss 25% of infected individuals, who are asymptomatic and could potentially be transmitting.

Overall, these findings suggest that aggressive contact-tracing strategies based on suspect cases may be appropriate early in an outbreak but as it progresses, control measures should transition to a combination of approaches that account for setting-specific transmission risk. Quarantine may need to cover entire communities such as dormitories, workplaces, or other institutional and residential settings, while contact tracing should shift to identifying hotspots of transmission and vulnerable populations. The variability in SAR across settings suggests targeted strategies may allow for reducing infections while not overly restricting social movement.

## Data Availability

Data used in the paper are from published articles and pre-prints. The compiled dataset can be made available upon request.

## Supplementary materials

**Figure S1a.**
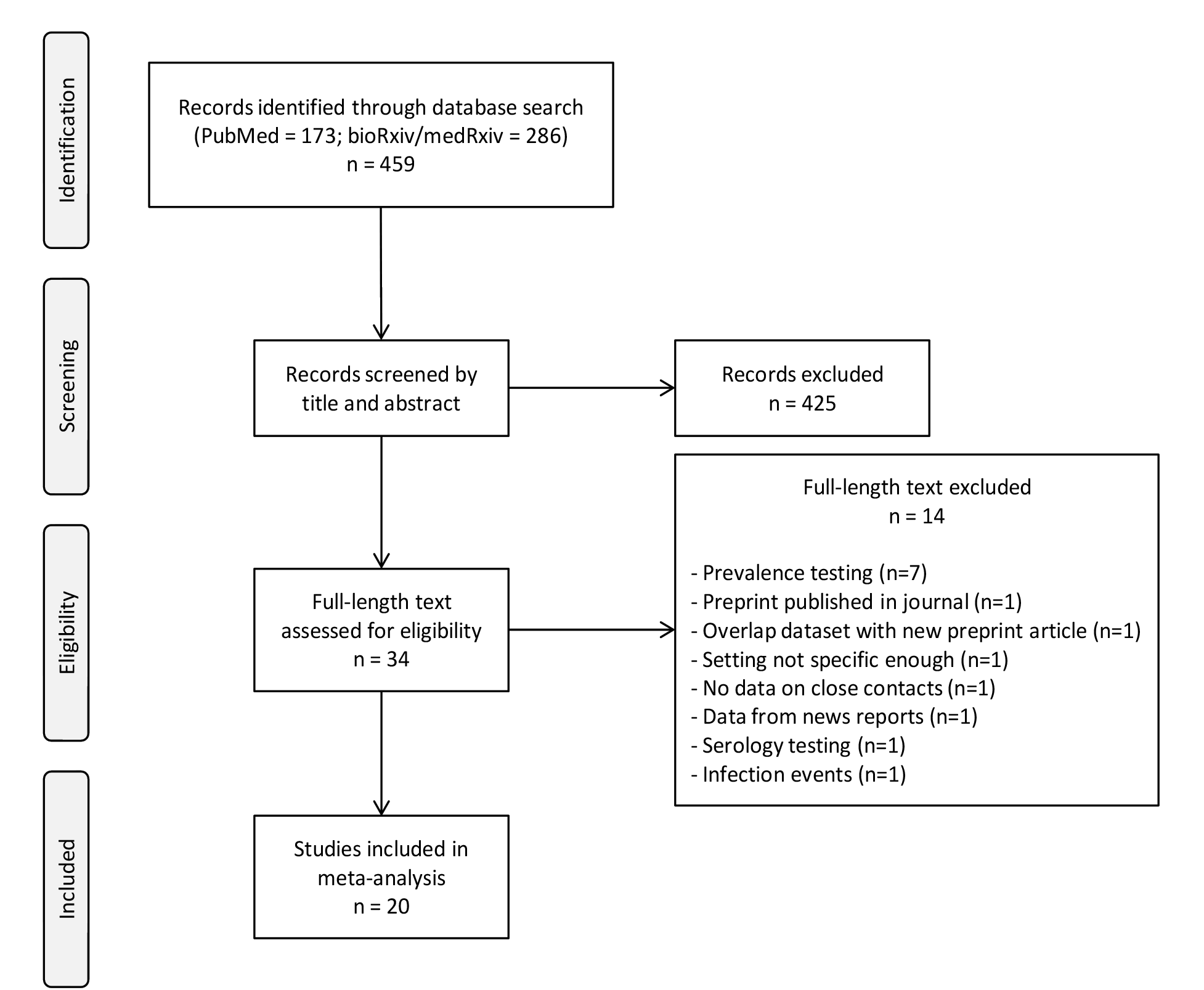
Flow chart of search strategy and study selection for the secondary attack rate (SAR)

**Figure S1b.**
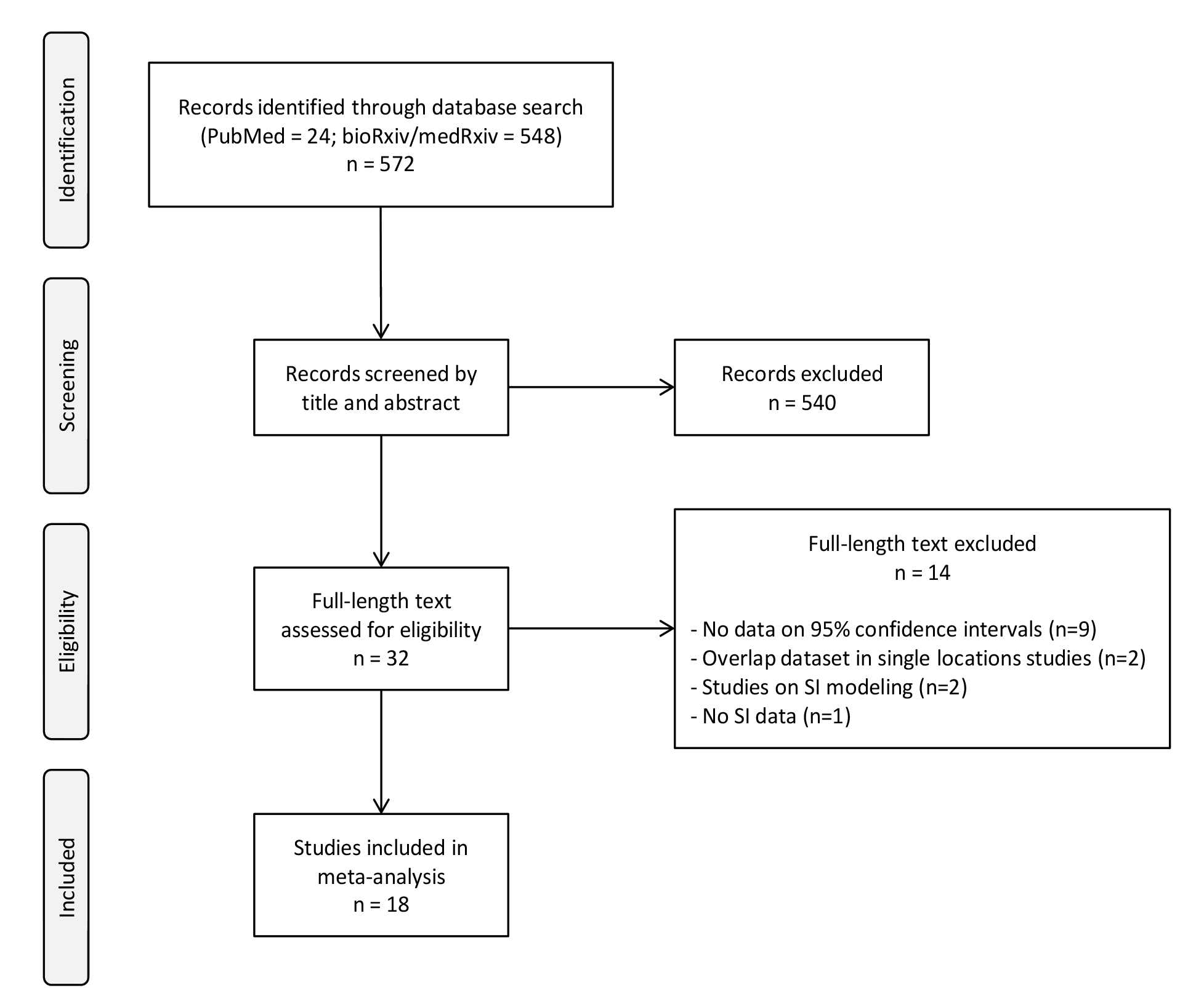
Flow chart of search strategy and study selection for the serial interval (SI)

**Figure S1c.**
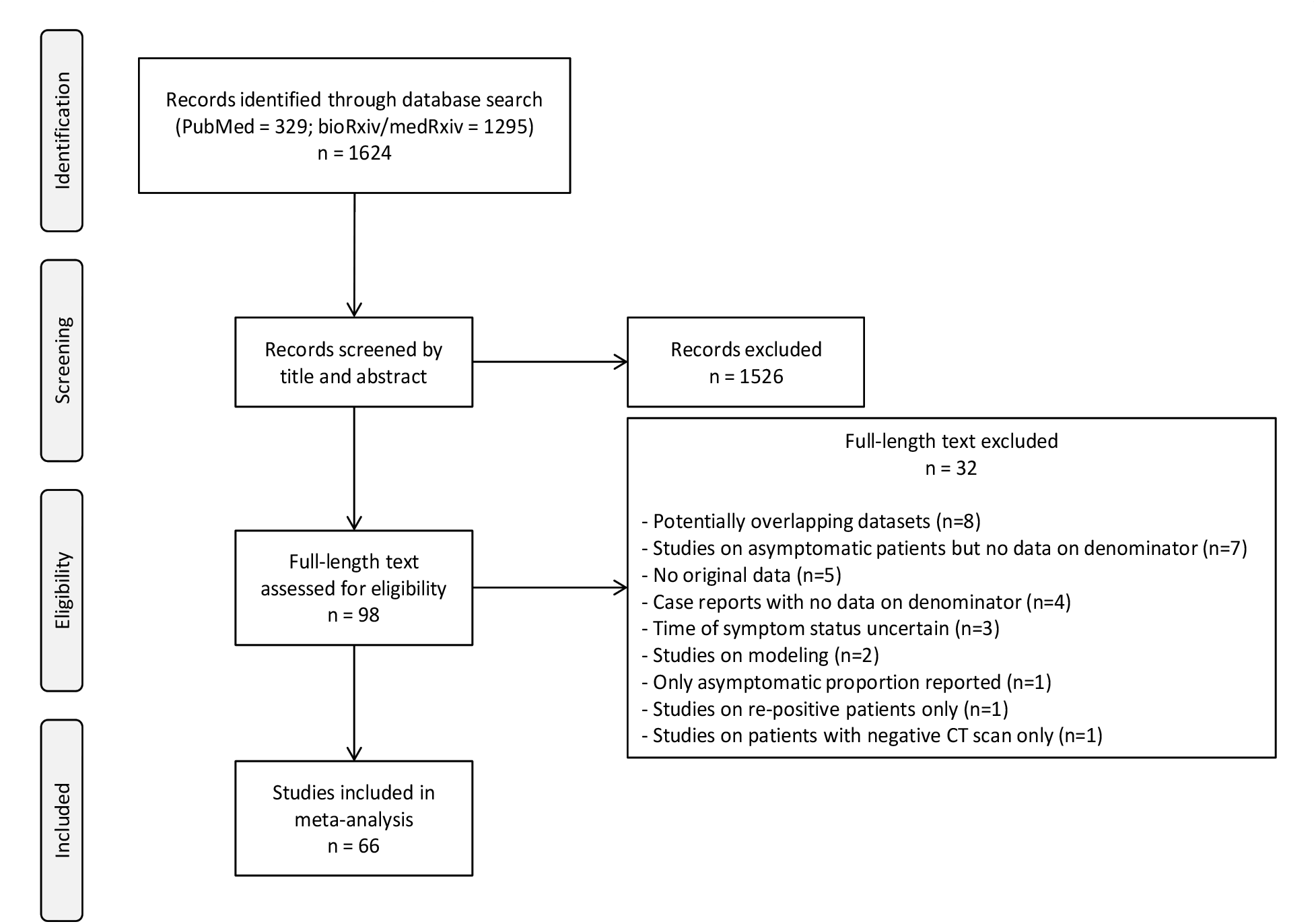
Flow chart of search strategy and study selection for asymptomatic infection.

**Figure S2a.**
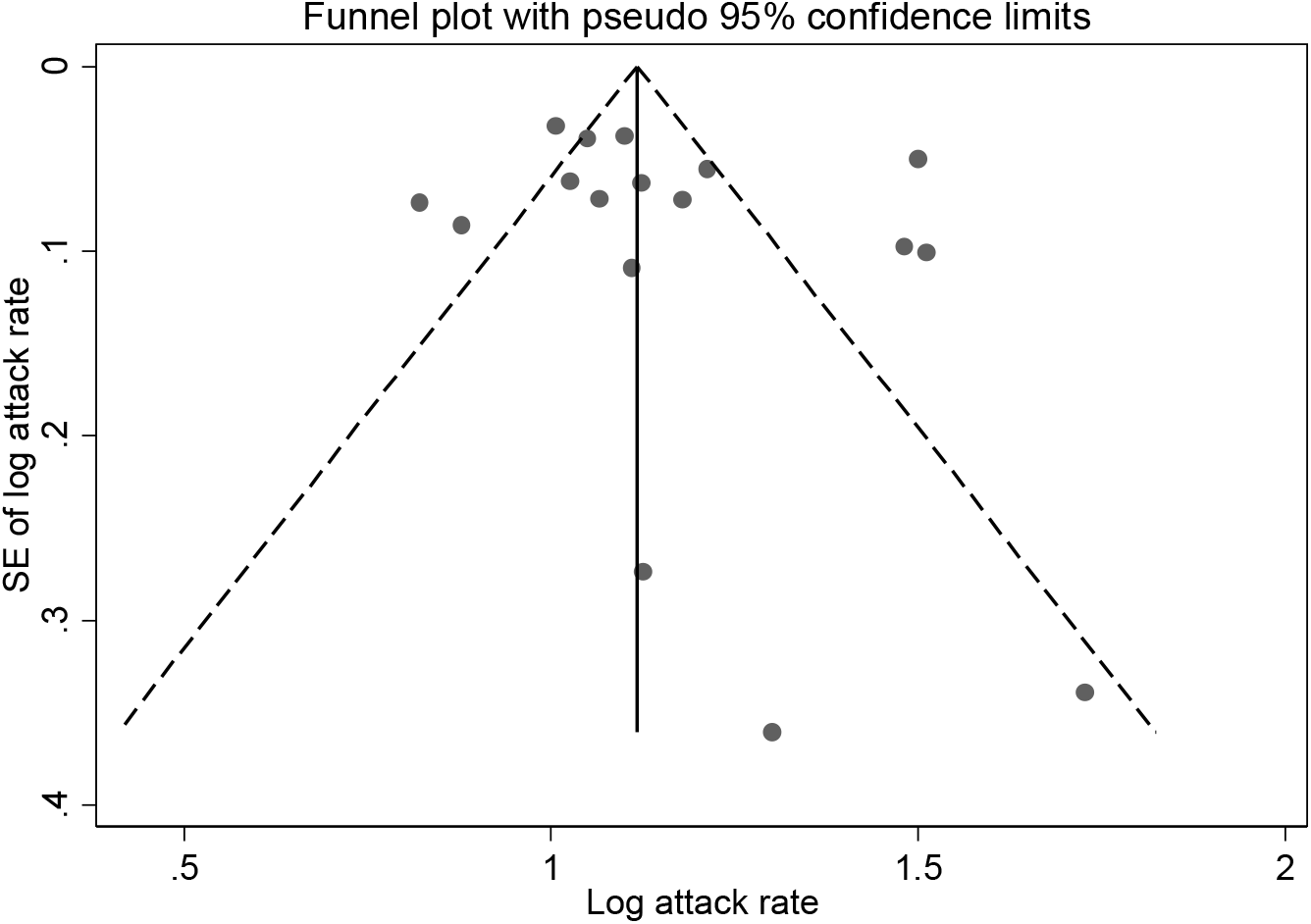
Funnel plot of the 17 studies on household secondary attack rate.

**Figure S2b.**
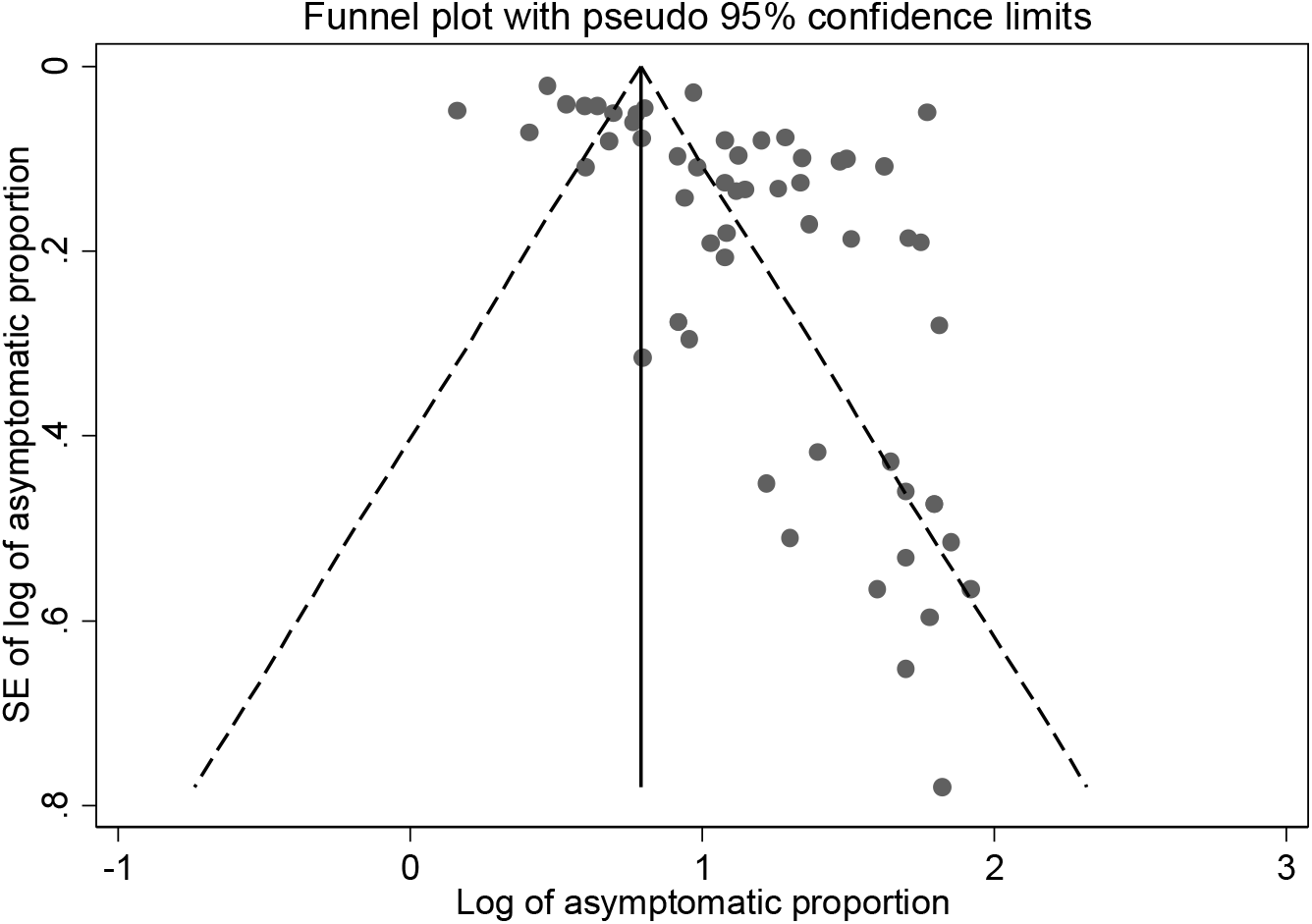
Funnel plot of the 57 studies on overall asymptomatic infection.

**Figure S2c.**
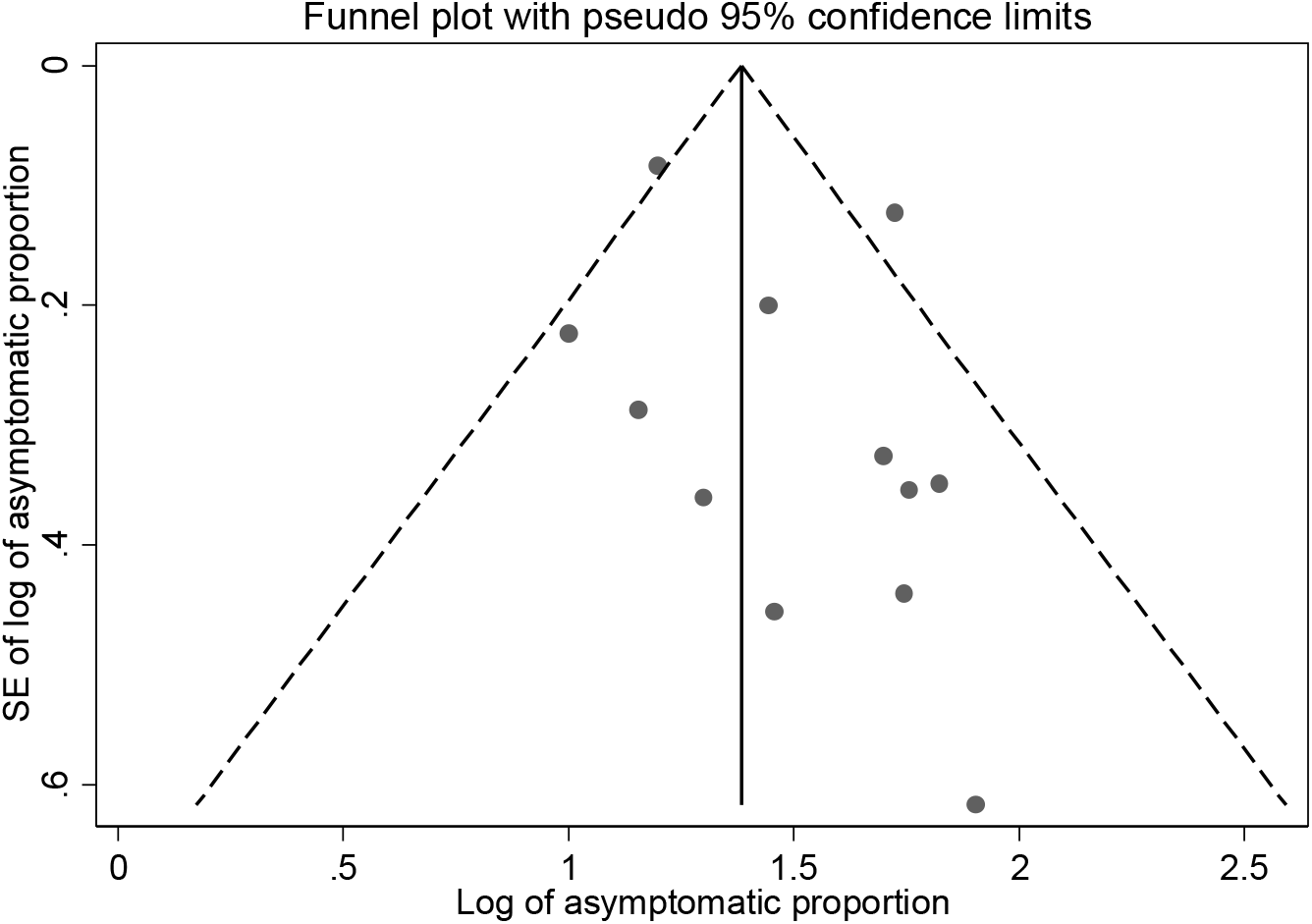
Funnel plot of the 12 studies on asymptomatic infection in children.

**Figure S3.**
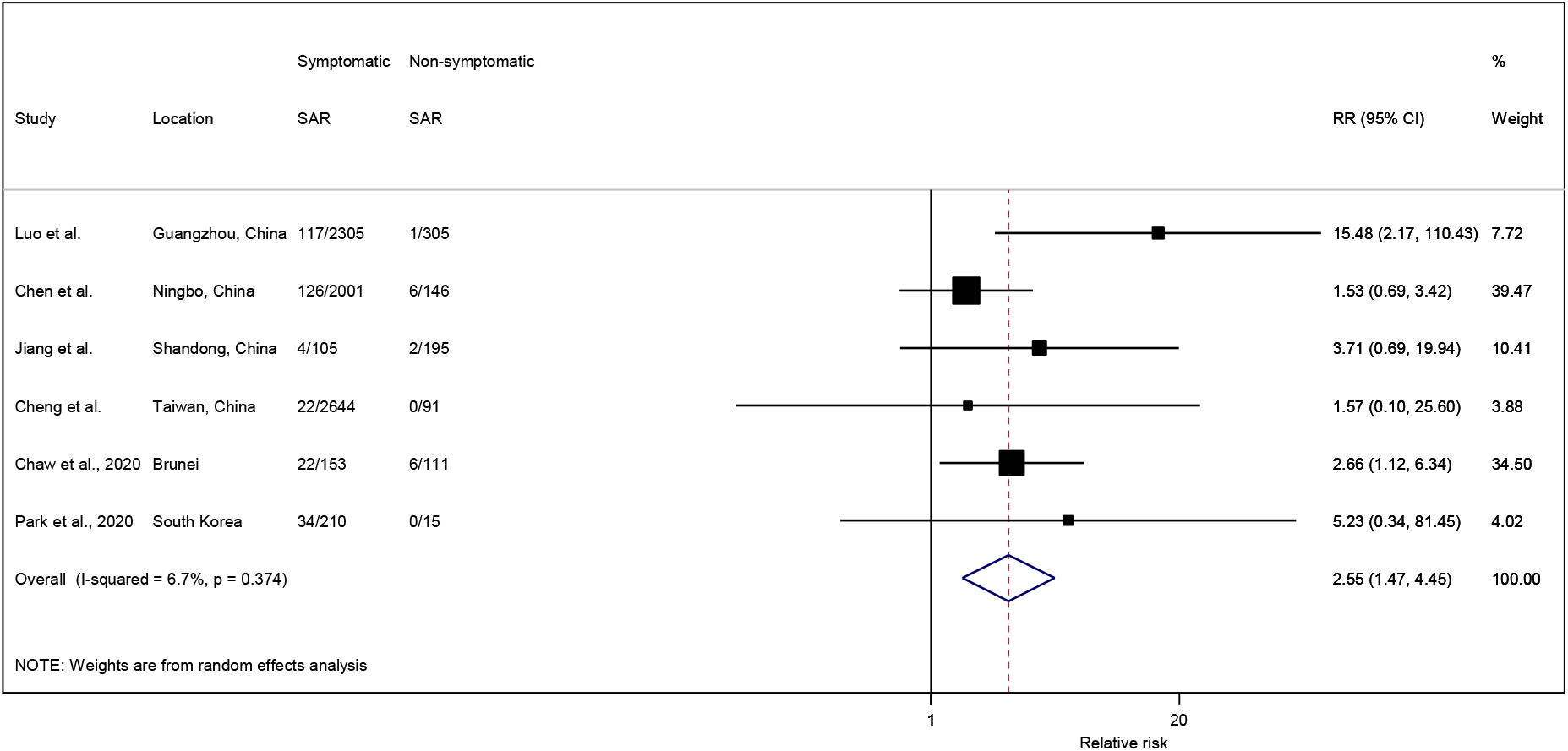
Forest plot of secondary attack rate by symptom status of index case. RR is the estimated relative risk ratio, with 95% confidence intervals (CI). I-squared is the percentage of between-study heterogeneity that is attributable to variability in the true effect, rather than sampling variation.

**Figure S4.**
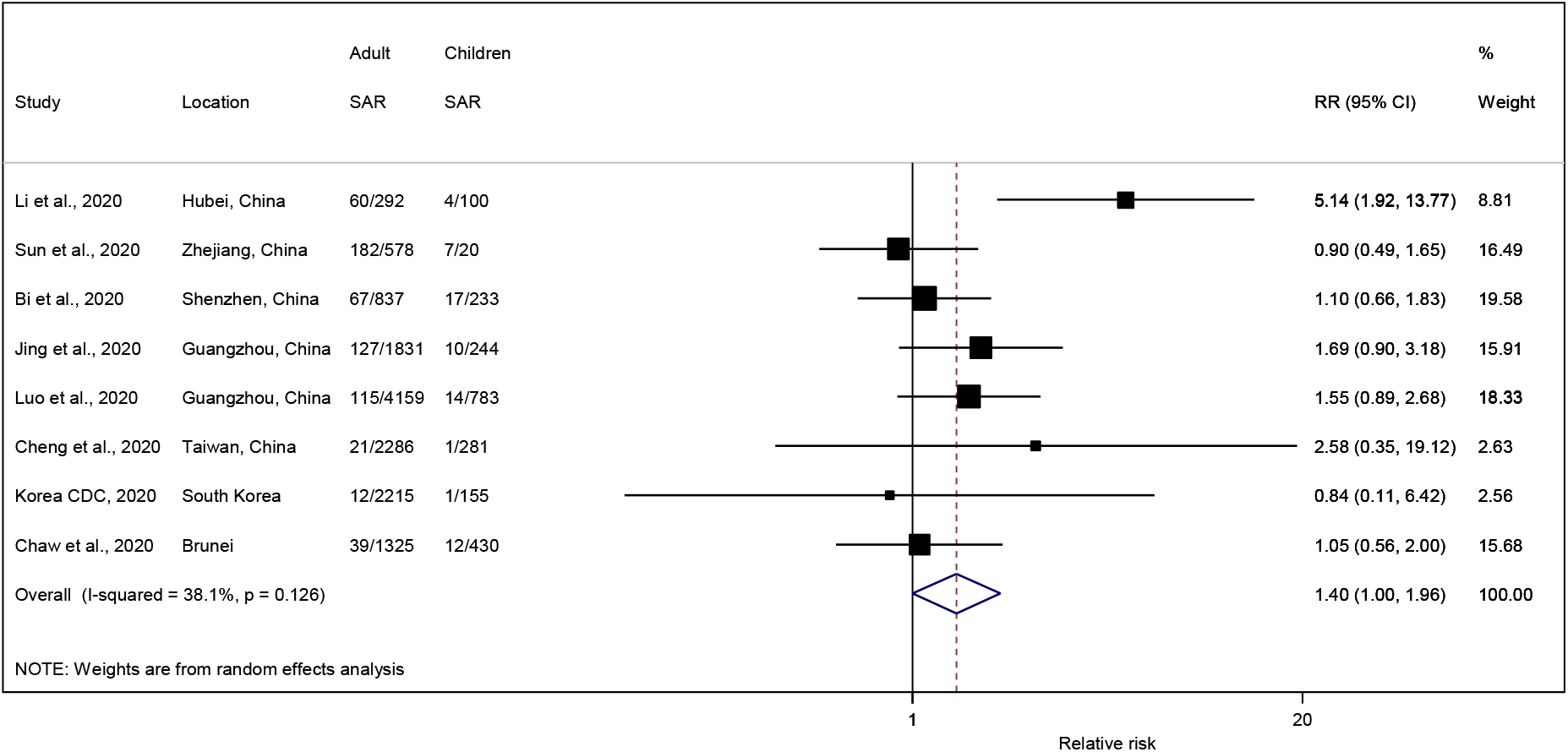
Forest plot of secondary attack rate in adults and children. RR is the estimated relative risk ratio, with 95% confidence intervals (CI). I-squared is the percentage of between-study heterogeneity that is attributable to variability in the true effect, rather than sampling variation.

**Figure S5.**
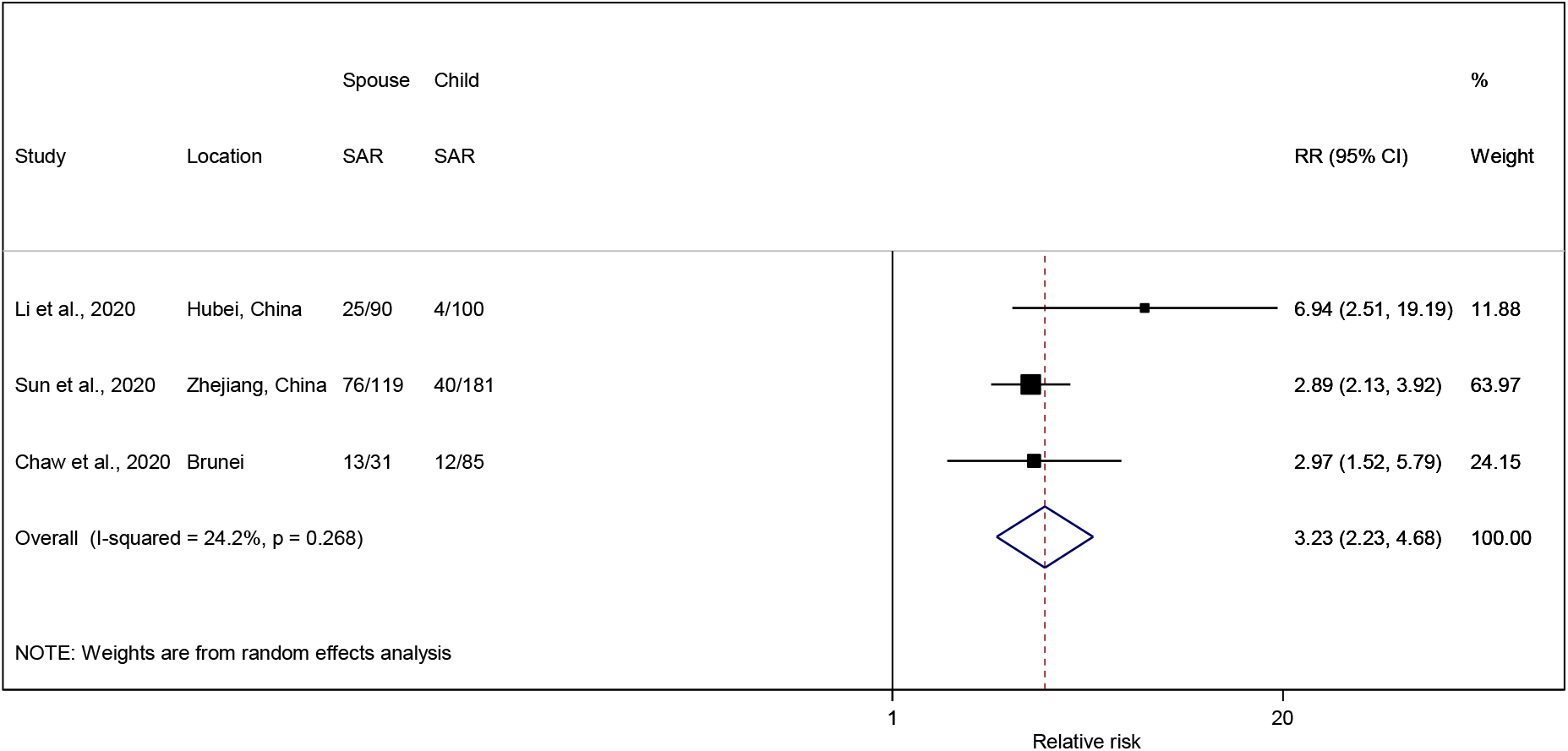
Forest plot of secondary attack rate in spouse and child of an index case. RR is the estimated relative risk ratio, with 95% confidence intervals (CI). I-squared is the percentage of between-study heterogeneity that is attributable to variability in the true effect, rather than sampling variation.

**Figure S6.**
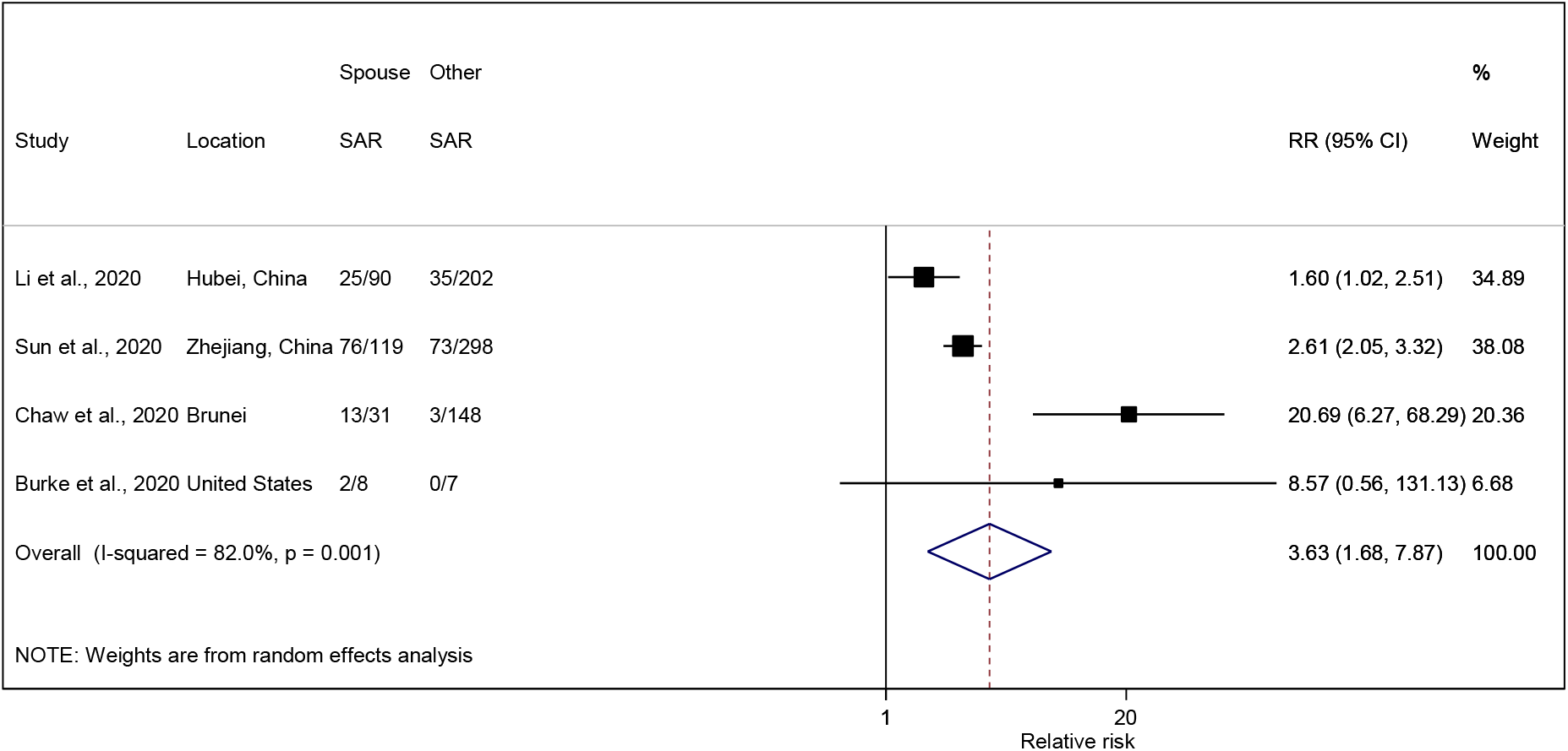
Forest plot of secondary attack rate in spouse and other household members (excluding child) of an index case. RR is the estimated relative risk ratio, with 95% confidence intervals (CI). I-squared is the percentage of between-study heterogeneity that is attributable to variability in the true effect, rather than sampling variation.

**Figure S7.**
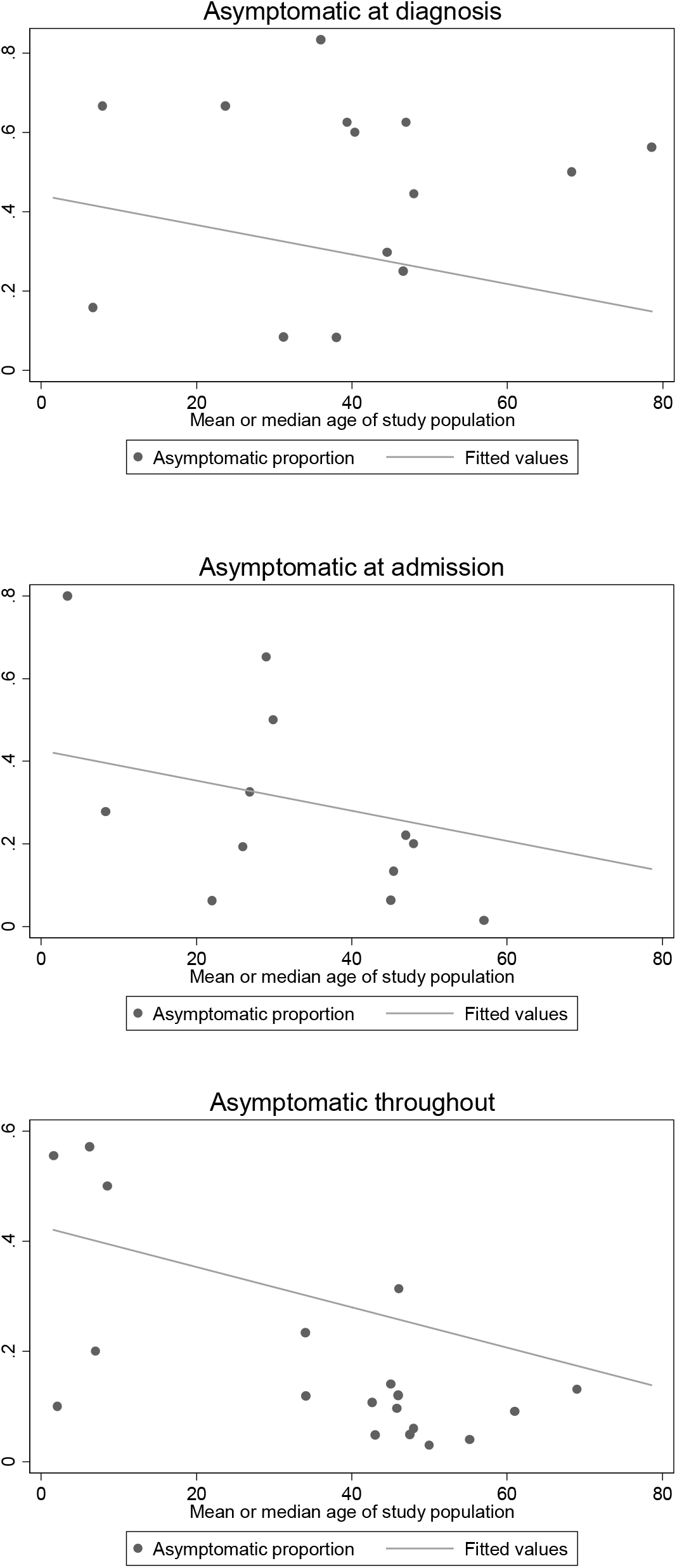
Scatter plots of asymptomatic infection and mean or median age of the study population.

**Figure S8.**
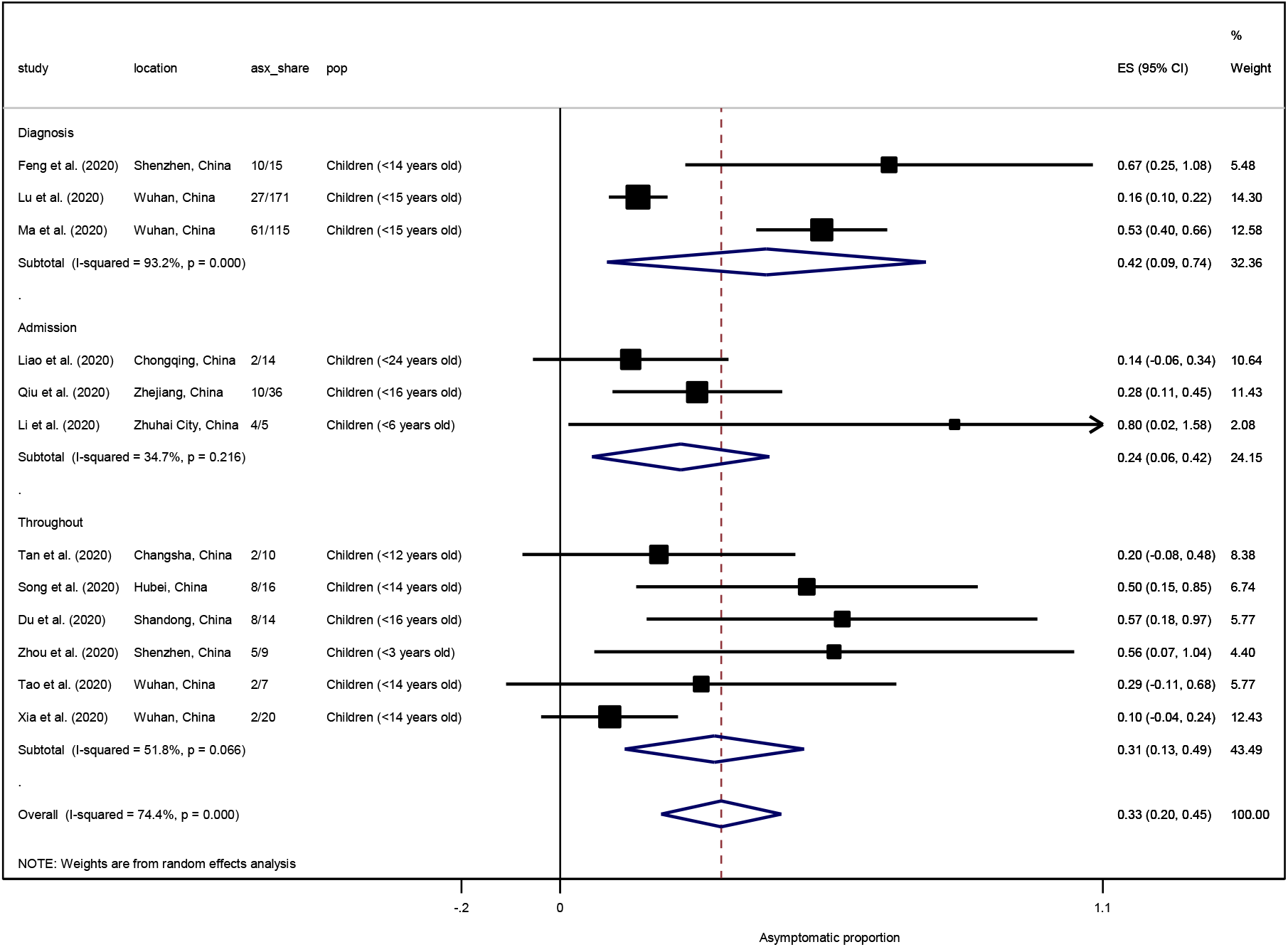
Forest plot of the proportion of asymptomatic cases in children at diagnosis, admission, and throughout the follow-up period. ES is the estimated asymptomatic proportion, with 95% confidence intervals (CI). I-squared is the percentage of between-study heterogeneity that is attributable to variability in the true effect, rather than sampling variation.

**Table S1a.** Results from Egger’s meta-regression test assessing the presence of small-study effects in 17 household secondary attack rate studies.

| Parameter | Estimate | SE | t value | p value | 95% CI |
| --- | --- | --- | --- | --- | --- |
| Slope (coefficient) | 1.038 | 0.090 | 11.48 | 0.000 | 0.846, 1.231 |
| Bias (intercept) | 1.464 | 1.479 | 0.99 | 0.338 | -1.689, 4.615 |

*Test of $H_0$ : no small-study effects, p value = 0.338*

**Table S1b.** Results from Egger’s meta-regression test assessing the presence of small-study effects in 57 studies on overall asymptomatic infection.

| Parameter | Estimate | SE | t value | p value | 95% CI |
| --- | --- | --- | --- | --- | --- |
| Slope (coefficient) | 0.597 | 0.067 | 8.98 | 0.000 | 0.464, 0.730 |
| Bias (intercept) | 3.414 | 0.870 | 3.92 | 0.000 | 1.670, 5.159 |

*Test of $H_0$ : no small-study effects, p value = 0.000*

**Table S1c.** Results from Egger’s meta-regression test assessing the presence of small-study effects in 12 studies on asymptomatic infection in children.

| Parameter | Estimate | SE | t value | p value | 95% CI |
| --- | --- | --- | --- | --- | --- |
| Slope (coefficient) | 1.243 | 0.138 | 9.04 | 0.000 | 0.937, 1.550 |
| Bias (intercept) | 0.862 | 0.702 | 1.23 | 0.247 | -0.701, 2.426 |

*Test of $H_0$ : no small-study effects, p value = 0.247*

**Table S1.** Attack rates in different settings.

| Study | Location | Setting | Attack rate (%) |
| --- | --- | --- | --- |
| Roxby et al. 2020 (68) | Seattle, Washington | Independent and assisted living community | 5/142 (3.5%) |
| Arons et al. 2020 (67) | King County, Washington | Nursing facility | 57/89 (64.0%) |
| Baggett et al. 2020 (142) | Boston | Homeless shelter | 147/408 (36.0%) |
| Ly et al. 2020 (70) | France | Homeless shelter | 48/683 (7.0%) |
| Lombardi et al. 2020 (66) | Lombardy, Italy | Healthcare workers | 138/1573 (8.8%) |
| James et al. 2020 (143) | Arkansas | Church | 35/92 (38.0%) |
| Park et al. 2020 (29) | Seoul, South Korea | Call center | 94/216 (43.5%) |
| Jang et al. 2020 (144) | Cheonan, South Korea | Fitness dance class | 57/217 (26.3%) |
| Dyal et al. 2020 (145) | United States | Meat processing plant | 4913/130578 (3.8%) |
| Zhang et al. 2020 (24) | Shandong, China | Supermarket | 11/120 (9.2%) |
| Yamahata et al. 2020 (65) | Japan | Cruise (Diamond Princess) | 696/3711 (18.8%) |

## Ethical approval

Not required.

## Authors’ contributions

JW and WCK conceived and designed the study. WCK, MAR, and MFA conducted the literature search and extracted the data. WCK performed the statistical analyses with input from LN. JW and WCK wrote the manuscript with critical feedback from MG, RP, LN, and LC. The final manuscript was approved by all the authors.

